# The Epidemiological Implications of Jails for Community, Corrections Officer, and Incarcerated Population Risks from COVID-19

**DOI:** 10.1101/2020.04.08.20058842

**Authors:** Eric Lofgren, Kristian Lum, Aaron Horowitz, Brooke Madubuonwu, Kellen Myers, Nina H. Fefferman

## Abstract

COVID-19 is challenging many societal institutions, including our criminal justice systems. Some have proposed or enacted (e.g. the State of New Jersey) reductions in the jail and/or prison populations. We present a mathematical model to explore the epidemiological impact of such interventions in jails and contrast them with the consequences of maintaining unaltered practices. We consider infection risk and likely in-custody deaths, and estimate how within-jail dynamics lead to spill-over risks, not only affecting incarcerated people, but increasing exposure, infection, and death rates for both corrections officers, and the broader community beyond the justice system. We show that, given a typical jail-community dynamic, operating in a business-as-usual way will result in significant and rapid loss of life. Large scale reductions in arrest and speeding of releases are likely to save the lives of incarcerated people, jail staff, and the community at large.

## Introduction

As the COVID-19 pandemic sweeps the globe, one of the critical functions of epidemiology is to consider how society can transform current practice to increase the health and safety of the public. Given the widespread risk of infection and the high case fatality rates, especially in older or medically compromised populations, the most effective strategies to reduce the impact of the disease may require that we be willing to consider structural reforms to our institutions to promote an overall greater good. To these ends, we have already seen systemic shifts in institutional practices that would be unthinkable under normal conditions: shelter-in-place orders closing businesses and restricting freedom of individual movement (*1*), school closures to limit transmission compromising the ongoing education of children (*2*), domestic travel restrictions and international border closures (*3*), suspension of visa processing (*4*), etc. Another clearly important institution that affects a substantial portion of the public directly (*5*) and an even greater portion indirectly (*6–9*), is our criminal legal system. The currently unfolding public health crisis makes clear the urgent need for rigorous analyses of the impact of maintaining current practices within these institutions, including both the costs to incarcerated people and their families as well as the costs to the community at large. We therefore explore the epidemiological costs associated with our current system’s functions as a necessary part of the policy conversation that must ensue to decide whether or not they should be maintained or altered in response to a growing global crisis, especially during the next few months as cases threaten to overwhelm healthcare systems throughout the United States. Already, some states have taken action to alter their incarceration practices to address the COVID-19 pandemic (e.g. NJ Senate Bill 2519, 2020), but many have not.

Analyzing incarcerated populations poses a unique epidemiological problem for several reasons. The population experiences high rates of movement and turnover (*10, 11*). Incarcerated people are responsible for purchasing their own hygiene products with limited resources (*12, 13*). It is difficult or impossible for incarcerated people to practice CDC recommended precautions for limiting transmission for a variety of factors including overcrowding and insufficient access to personal protective equipment (*14–20*). The incarcerated population has a higher expected rate of existing health conditions than the community from which they come (*21–23*). Jails are dependent completely on a workforce that moves in and out of the jail and the community including vendors, lawyers, corrections officers, medical staff, etc. And, there is strong evidence that incarceration itself has profound adverse effects on the health of incarcerated people (*24–26*). These descriptors make jails highly likely not only to place detained people at increased risk of infection and resulting severe outcomes, but also to function as a driver for increased infectivity, adversely impacting attempts to contain and mitigate disease spread in the broader communities in which jails are located.

To study the dynamics of this system and provide quantitative metrics for risk to incarcerated populations and the populations with which incarcerated people necessarily interact, we construct and tailor a epidemiological model of COVID-19 transmission, and then use that model to consider how some possible reforms to the system will alter these baseline risks. In doing so, we focus only on interventions that do not rely on the suspension of any individual rights guaranteed to individuals by either the US or individual state constitutions, but instead rely on elements of the criminal justice system that are already at the discretion of law enforcement, departments of corrections, and the court system (i.e. reduction in arrest intake, increased rates of returning incarcerated people to their homes, and improvement of conditions within the jails-indeed, these actions have already been undertaken by isolated, individual jails). We parameterize our model with data available from the Allegheny county jail system and perform a broad sensitivity analysis and comparison of relevant metrics to demonstrate how such models may apply to jail systems throughout the United States.

## Results

Unsurprisingly given the epidemiological dynamics of COVID-19, absent any intervention there is a substantial outbreak in the community, causing 1,051,238 infections (both symptomatic and asymptomatic) as well as requiring 71,735 hospitalizations and ultimately resulting in 11,203 fatalities over the 180 days of the simulation, with the peak of infections occurring 81 days after the first infective case appeared in the population. Among those incarcerated, the outbreak is considerably more severe, causing a cumulative 4,779 cases requiring 312 hospitalizations and 58 deaths among those incarcerated, the 2500 person jail being 0.2% the size of the wider community (Fig 1). The peak of this within-jail epidemic is also considerably earlier, with the peak of the epidemic occurring 29 days after the first infective case appeared in the community. Given the dominant approach to controlling COVID-19 in the community and the widespread calls to “flatten the curve,” for the remaining results we assume the presence of a shelter-in-place order or similar social distancing intervention *only in the community* as the comparator scenario, represented as a reduction in the mixing frequency of all age groups in the community. In line with the experience of communities undergoing such distancing interventions, this decrease in overall contacts results in a substantially delayed epidemic, with 39,965 infections in the community as well as far lower burdens in terms of both hospitalizations and fatalities. In contrast, the early dynamics of the COVID-19 outbreak within the incarcerated population are identical, while in the latter half of the simulation the outbreak dynamics in the incarcerated population are markedly worse, resulting in 8,339 infections after 180 days and proportionately more hospitalizations and COVID-19 related fatalities (Fig 2). Shelter-in-place orders had no discernible impact on the health outcomes of the staff of the jail.

**Fig. 1.**
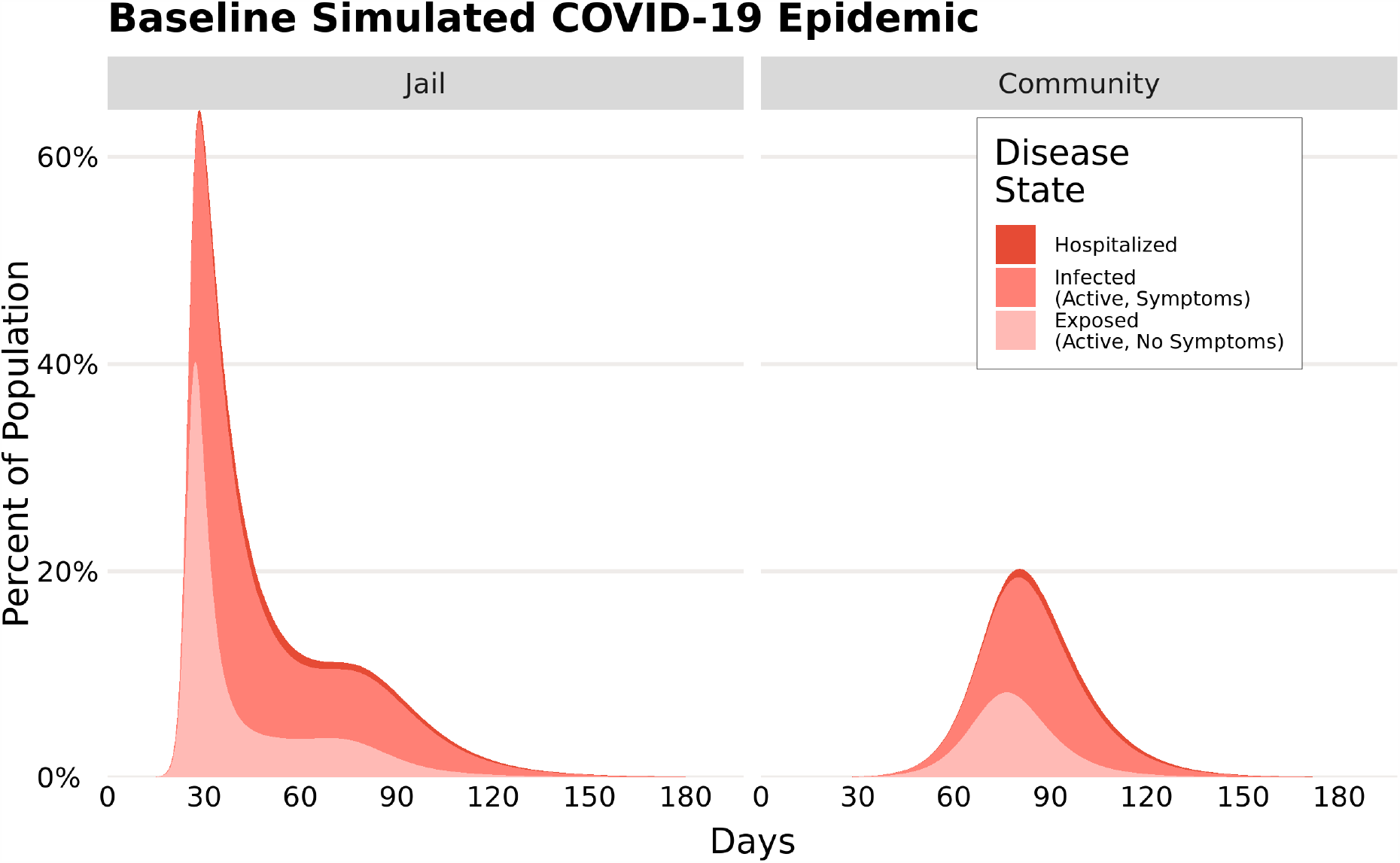
Epidemic curves from a simulated COVID-19 epidemic in an urban community (right) and the connected population of persons in a jail (left). The curves demonstrate the expected magnitude and timing of the outbreak in the different populations, broken into the different etiologically relevant categories (i.e. Exposed, Infected, and Hospitalized).

**Fig. 2.**
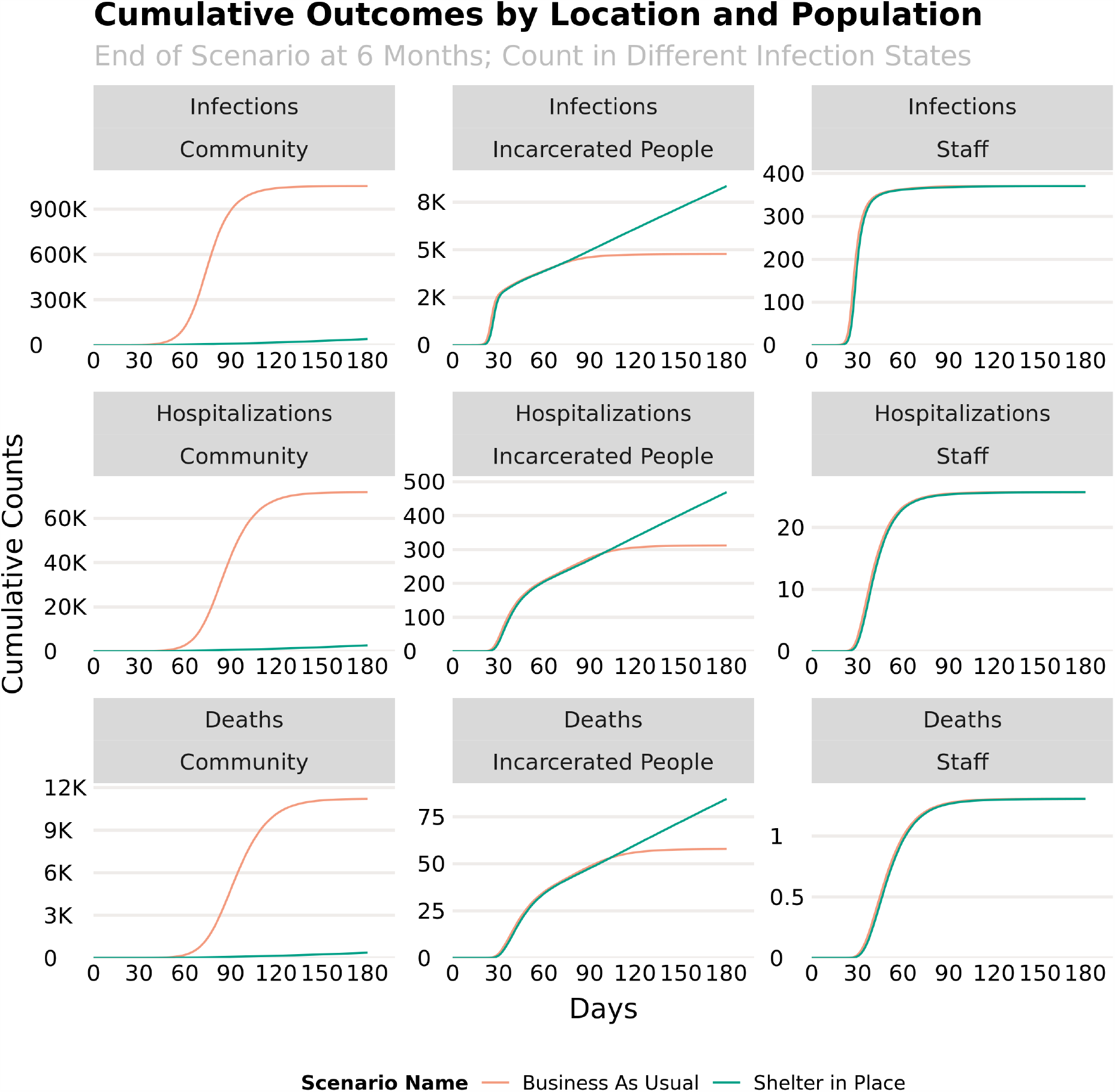
The impact of social distancing on cumulative outcomes for each population. Cumulative infections, hospitalizations and deaths reveal different dynamics in the community (first column), among persons in jail (second column) and among jail staff (third column) for scenarios with (green) and without (orange) a shelter-in-place social distancing intervention.

All of the four considered arrest deferral scenarios had substantial impacts on the course of the epidemic in the incarcerated population, while also lessening the impact of the epidemic on the community and, to a lesser extent, the jail’s staff. Discontinuing the arrest of bail-eligible individuals, which corresponds to a *≈*25% reduction of admissions into the jail, resulted in a 24% reduction in infections in the incarcerated population, and a 20% reduction in infections within the community. Note that, as discussed above, this model seeks to examine the role of jails in a population otherwise doing relatively well in controlling their outbreak. As such, these numbers arise in a population with a sustained, six-month, broad-based shelter-in-place order, and as such the cases arising from the jail population as a percentage of total cases are expected to be higher than they would be in a population with less stringent controls.

Broader, more sweeping arrest deferral programs resulted in correspondingly larger impacts in both the incarcerated population and the community as a whole. The discontinuation of arresting individuals for low level offenses (*≈*83.4% reduction) and the blanket reduction of arrests by 90% resulted in a 77% and 80% reduction in infections within the incarcerated population (with correspondingly fewer hospitalizations and deaths) respectively. These strategies also resulted in the greatest decrease in infections among staff (11% and 14%) and in the community at large (62% and 66%). Finally, a strategy built on deferring the arrest of individuals at high risk of developing COVID-19 related complications by 90%, tailoring the intervention to groups of epidemiological importance rather than the nature of their offense, resulted in a 30% decrease in infections within the incarcerated population and a 61% decrease in deaths among the same population (Fig 3).

**Fig. 3.**
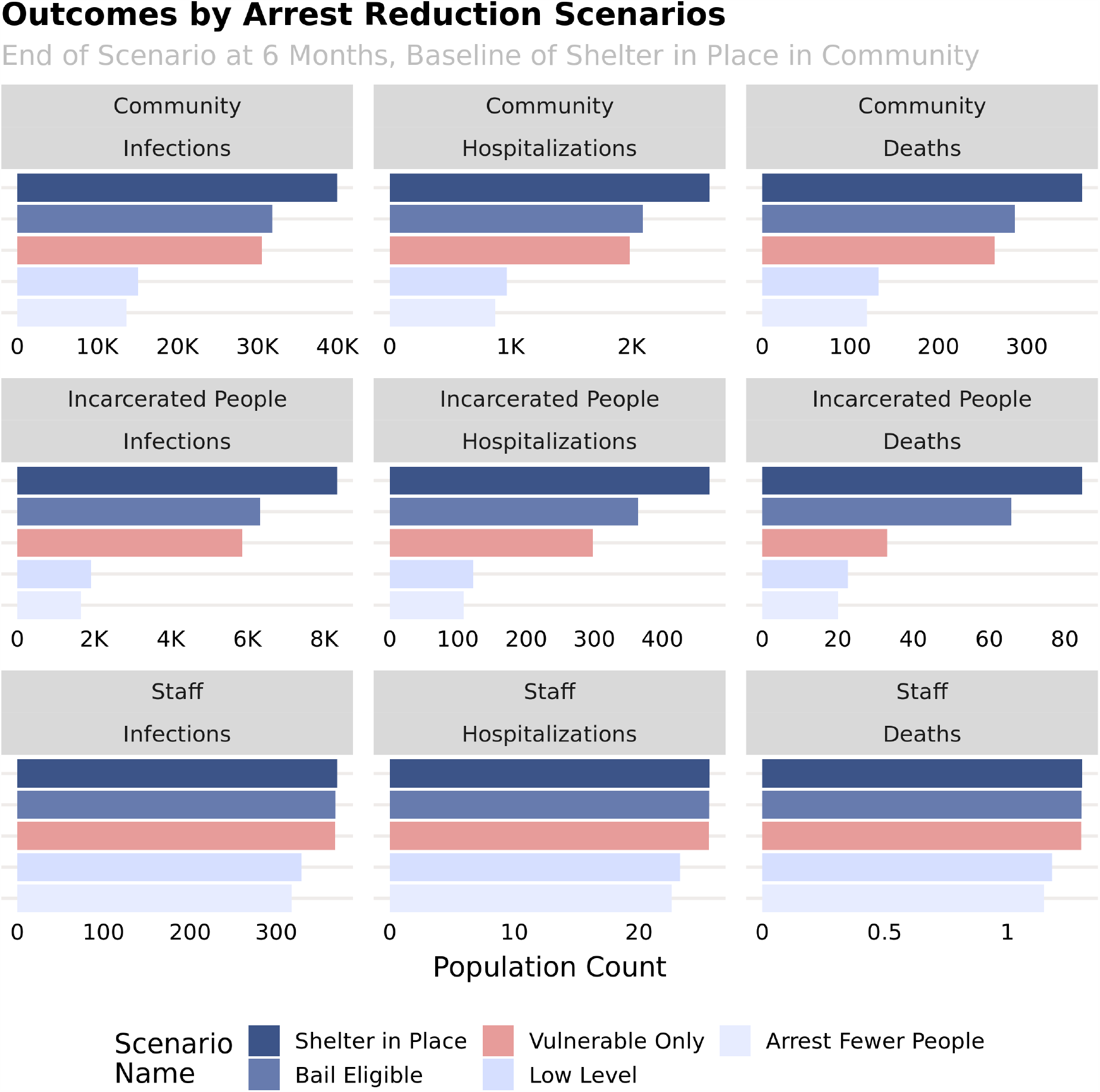
Cumulative infections, hospitalizations and deaths in the community (first row), among persons in jail (second row) and among jail staff (third row) for several incarceration deferment scenarios.

In comparison, a strategy deferring the same number of people with no regard to their underlying risk, had smaller effects on the number of infections and deaths. Specifically, releasing the same number of individuals as in the above scenario without regard to their risk resulted in only a 19% decrease in infections within the incarcerated population and a 18% reduction in deaths. The deferral strategy targeting individuals for high risk outcomes caused 0.9% more infections in the community relative to the same proportionately large but broader strategy, and the decreased number of deaths among persons in jail was partially offset by this increase. The targeted strategy resulted in a combined number of COVID-19 fatalities in both the population of persons in jail and in the community of 293 compared to 372 fatalities under the broader strategy.

Pairing increased arrest deferral with a more rapid release of persons who were already incarcerated enhanced the impact of those interventions, reducing infections, hospitalizations and deaths overall (Fig 4). The rate of decrease was less dramatic in the community and staff populations, especially at lower levels of accelerated release schedules.

**Fig. 4.**
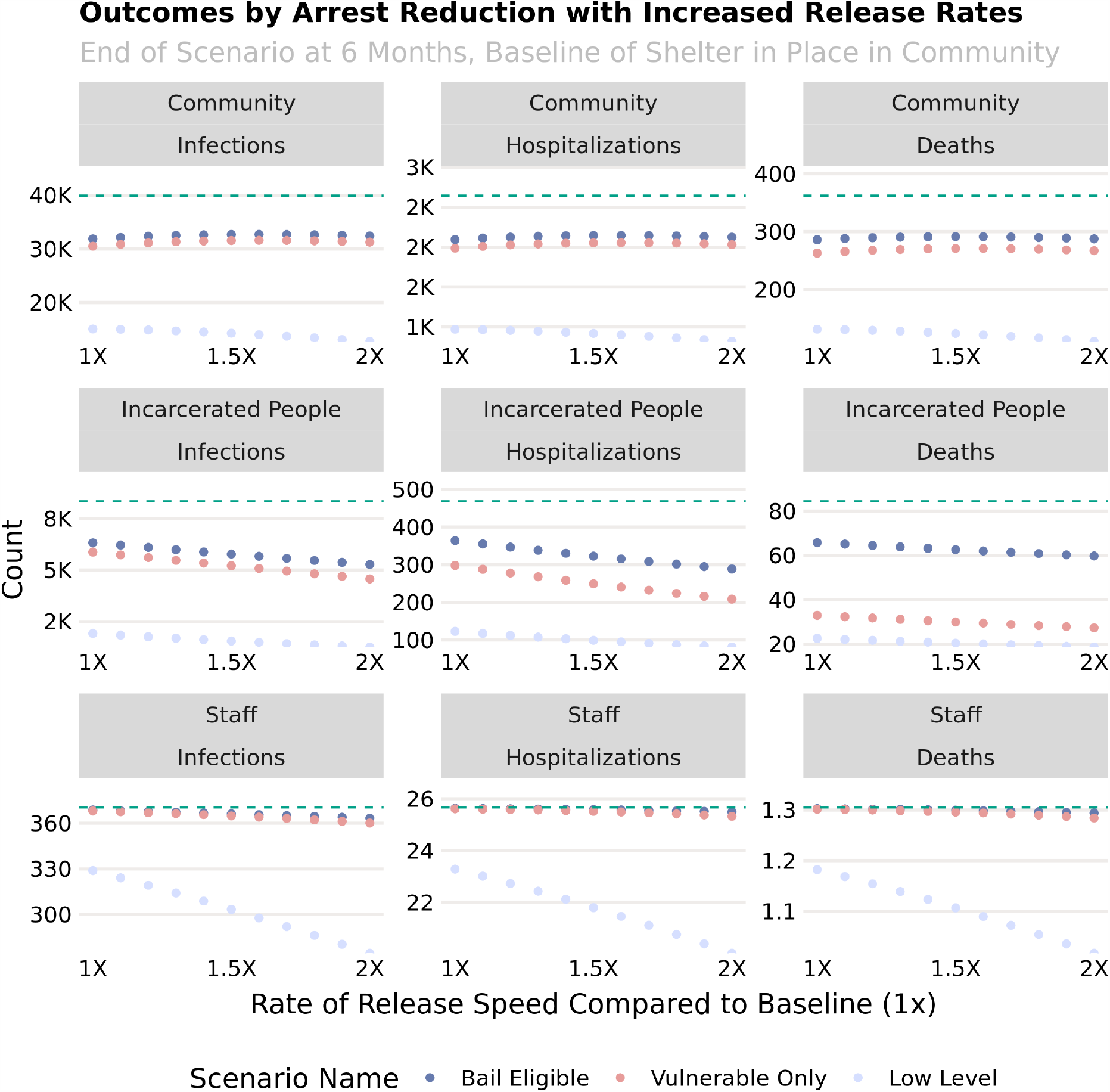
Cumulative infections, hospitalizations and deaths in the community (first row), among persons in jail (second row) and among jail staff (third row) for several combinations of incarceration deferment and accelerated release.

When accompanied by the deferred arrest of bail-eligible individuals to reduce the incarcerated population, supplementary measures to reduce transmission among incarcerated persons have a marked benefit in both reducing the amplitude of the epidemic curve in incarcerated people and jail staff, as well as shifting the overall community epidemic curve later (Fig 5). These interventions may be thought of as either measures to reduce mixing — such as allowing greater space between individuals in common areas or the staggering of the use of shared facilities — or the provision of supplies such as soap and hand sanitizer that reduces the level of viral contamination of patient’s hands, physical surfaces, etc. Compared to the baseline mixing rate among persons in jail, a reduction to an equivalent level of mixing as the community while sheltering in place would reduce infections in this population by 37% as well as delay the peak of the epidemic by approximately 40 days.

**Fig. 5.**
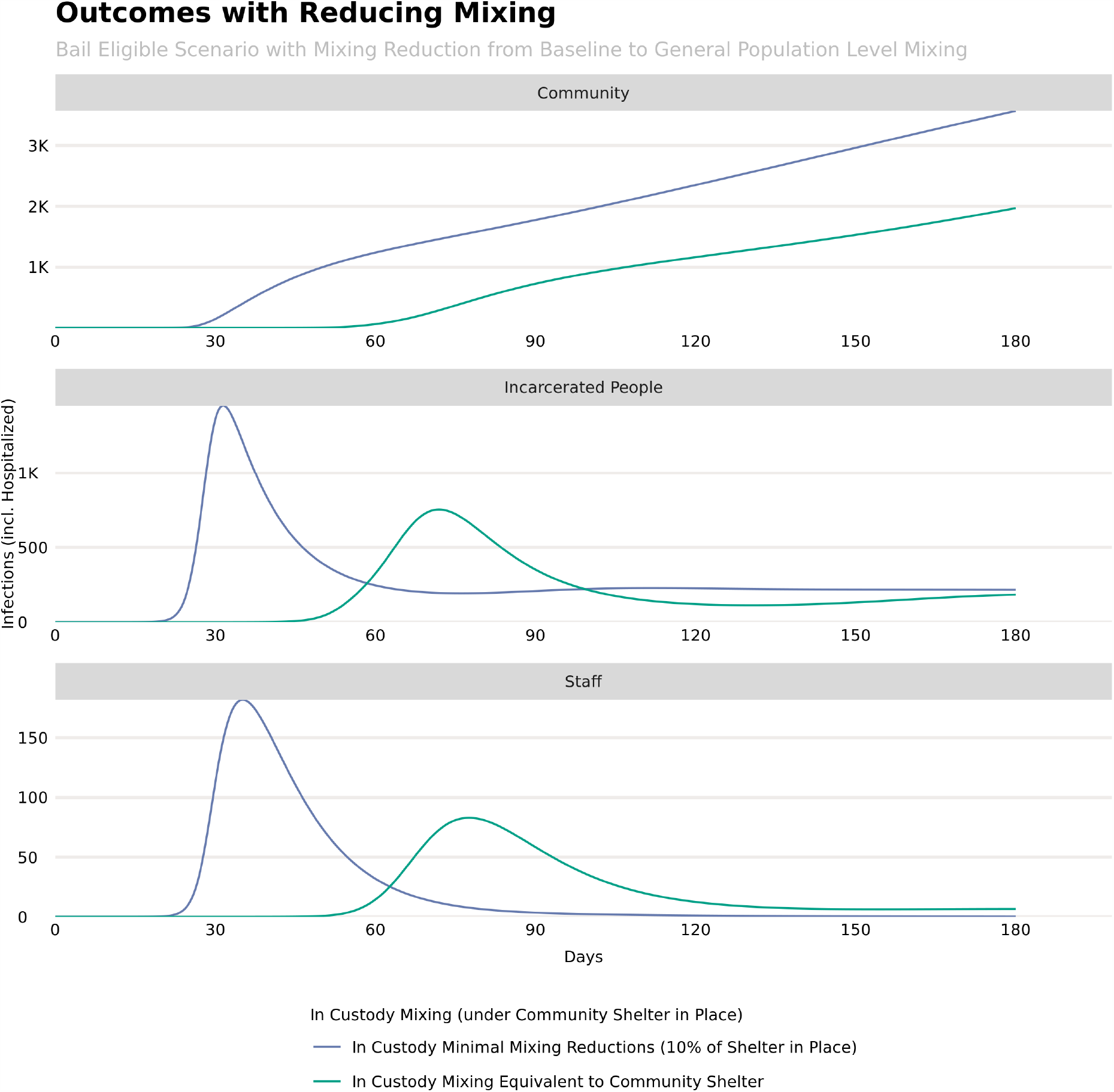
Epidemic curves for the community (top panel), persons in jail (middle panel) and jail staff (bottom panel) under a shelter in place order as well as the deferment of bail-eligible persons. The curves show the impact of increased reduction in mixing (e.g. from the ability to physically distance persons in jail while in common areas) from baseline (dark blue) to identical to the community’s shelter-in-place order (green).

An increase in the detection of severe COVID-19 cases among incarcerated persons from 95% to 100% (equivalent to the same detection of the need for medical treatment available in the community) unsurprisingly increased the number of hospitalizations, as 5 out of every 100 incarcerated persons needing hospitalization were no longer missed, either for lack of access to care, insufficient diagnostic capacity, or other reasons. Similarly, owing to the vast reduction in the case fatality rate between hospitalized (CFR = 5% for low risk and 33.3% for high risk) severe cases and unhospitalized severe cases (CFR = 100% for both groups), the number of deaths dropped by 91% when the detection of severe cases rose to the same level as the community. Between these scenarios, the number of infections rose slightly with better detection, increasing by 0.2% (Fig 6). This is likely due to the slightly longer time an untreated severe case spends in the incarcerated population before they are removed due to death vs. when a treated case is transferred for hospitalization. This effect will only be present if the level of viral shedding is constant (or increasing) over the course of a clinical infection. If instead the mechanism by which a severe COVID-19 patient dies is a cytokine storm or other process not involving the virus overwhelming the immune system, we would not expect this effect to be observed. However, even in the pessimistic case wherein viral shedding is constant throughout the clinical course of infection, the slight rise in infections is offset by the decrease in the number of COVID-19 related fatalities.

**Fig. 6.**
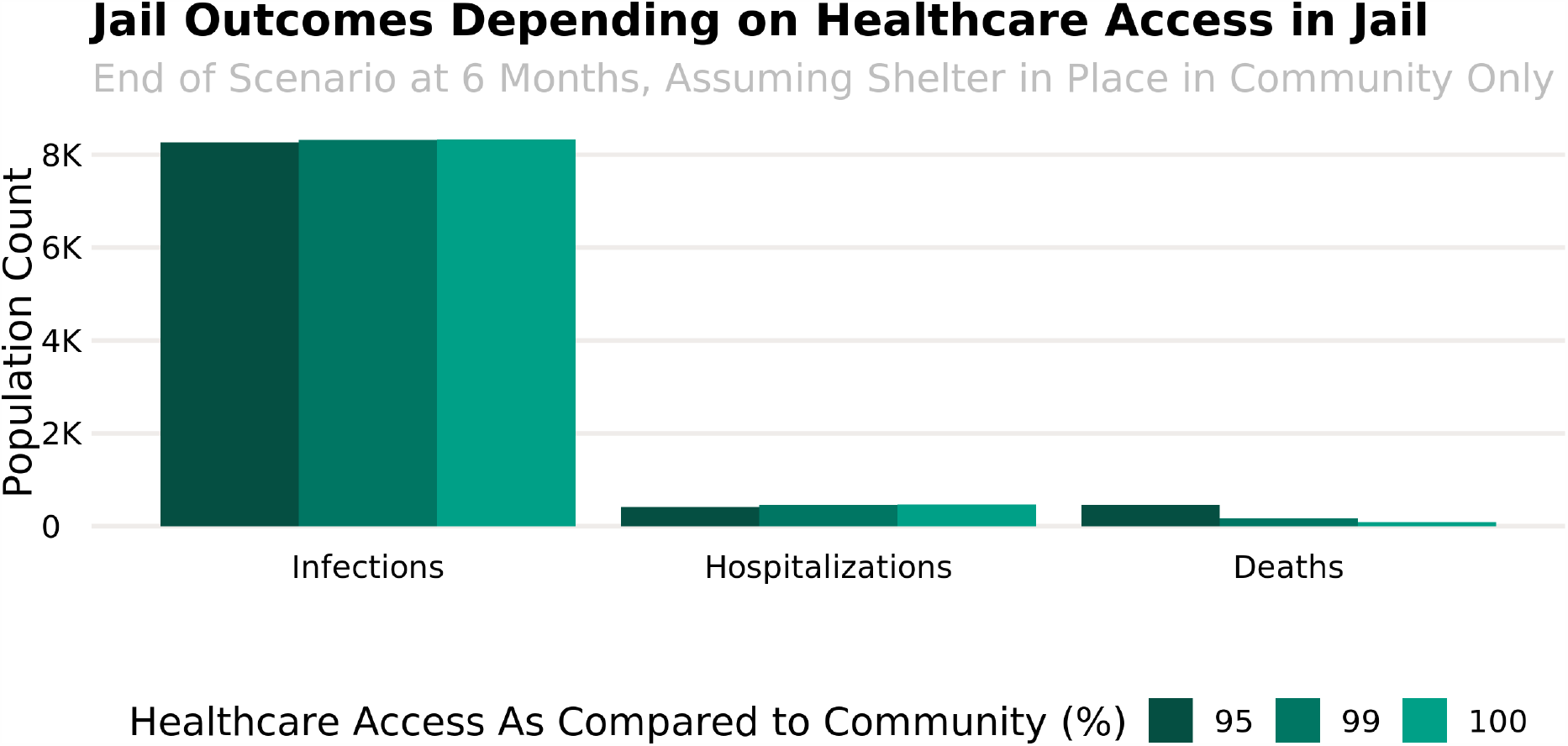
Infections, hospitalizations and deaths from COVID-19 among incarcerated persons under three scenarios of the probability that a severe case will be detected and receive adequate medical care.

## Discussion

Failure to adequately protect jail populations will have a profound impact on the health of incarcerated people, corrections officers, workers in the judicial system, and members of the broader community. Our model demonstrates that, in the absence of community mitigation such as strict social distancing, by only 30 days after the introduction of the first infection to the community, we can expect 2628 infections among incarcerated people, resulting in two in-custody deaths. These results clearly follow from the features of the jail system themselves in challenging ways. While only 1% of the population entering into the jail system are elderly (*27*), incarceration in jail itself degrades the health of incarcerated people (*24–26*), leaving them more vulnerable to infection and severe outcomes from infection (*28*). As individual robustness to disease decreases, the epidemiological result is the increased vulnerability of the whole jail population.

Beyond the direct implications for the health of incarcerated people, jail populations have high rates of re-entry into the general community and they depend on people who regularly mix with the outside community. It is worth noting that, even as some court systems transitioned to video-conferencing and remote hearings, incarcerated populations do not have access to tele-conferencing capabilities where they are housed, and must still be transported to, and staffed while using, communications facilities. Jail populations are largely composed of individuals who have not been convicted of a crime, and therefore will be released quickly back into the general community rather than to further incarceration within the carceral system. Jails with disease prevalence higher than the general populations they serve will therefore act as sources of infection, re-seeding infection into communities that may be striving to contain or mitigate ongoing outbreaks, or even reintroducing infection into otherwise disease-free populations. Dynamics consistent with these predictions have already been observed (*29,30*), but can and should be considered as an ongoing challenge. It is important to note that this would happen even if no one were released given the volume of people coming in and out of jails in staff and vendor roles, so should therefore not be construed as an implication that releases should be suspended or impeded. As COVID-19 continues to spread throughout the US, tracking data within jails and the communities they serve will be critical in validating studies such as this one and in shaping best practices to limit jail-driven spread going forwards.

We are not helpless to effect change. Some obvious potential courses of action suggest themselves immediately. New arrests mean that people of unknown disease status may be regularly brought into jails, increasing the likely severity of outbreaks both by the plausible continuous introduction of new sources of infection and by the maintenance of higher rates of contact among susceptible incarcerated people due to the density and structure of jail housing arrangements. If jurisdictions across the country reduce their intake by significant percentages, our models demonstrate that we will meaningfully directly reduce the disease incidence in the incarcerated population (as seen in Fig 3). Moreover, these same strategies also clearly produced a reduction in the source of risk to incarcerated people’s families, jail staff, and the broader community (Fig 3). These strategies could be enacted in a number of ways, such as (but not limited to) replacing misdemeanor arrests with citations, avoiding recommendations for jail time or prohibitive terms for bail conditions, or refusing to detain anyone for nonpayment of fines or fees during the course of the outbreak.

Having considered these potential strategies for categorical reduction in intake into jails, we also considered the case in which the categorical consideration for reduction in intake stemmed instead from the health of the arrested person. While this results in a smaller within-jail outbreak and reduces the resulting fatalities, we fail we fail to achieve any significant reduction in disease burden in the broader community by taking this action. It is therefore more effective to more aggressively reduce the intake rate across the entire population than to attempt to single out particular categories of individuals due to their likely susceptibility to severe morbidity or mortality from infection. The larger the reduction in overall intake, the greater the reduction in disease achieved for all populations (incarcerated people, the broader community, and jail staff, in decreasing proportion of effect). These broader interventions are also likely to be relatively straightforward to implement administratively, without knowledge of an individual’s underlying comorbidities, if any.

In addition to reducing rates of intake into the jail system, another obvious, concrete step we might take to reduce disease risks for everyone is to increase the rate of release from jails. This should clearly be coupled with a decreased rate of intake rather than enacted in isolation, since increasing release rates while maintaining the same rate of intake would increase infection risks for incarcerated people, the staff who work at the jails and court systems, and the broader community. This may even still occur when expedited release is coupled with decreased rates of intake if the rate of release is insufficient; see Fig 4. Again, our results clearly demonstrate that the greater the proportion of the incarcerated population we can include in such a policy, the more effective the intervention is at mitigating the outbreak.

To be maximally effective, each of these interventions should anticipate, rather than react to, widespread infection incidence in jail populations.

Critically, the factors that cause these outbreak dynamics and drive the resulting efficacy of proposed interventions are features implicit in the nature of the jail system itself. The living conditions foster disease spread. Incarcerated people are shuttled back and forth to court or, where court proceedings are halted due to this pandemic, forced to remain in their cells or dorms. Incarcerated people occupy shared spaces in which physical distancing is impossible either due to space, overcrowding, or the requirement of constant supervision. Incarcerated people are often not provided with the means to disinfect their surroundings or practice all of the hygiene guidelines suggested by the CDC. Improved facility sanitation, access to free personal hygienic care, such as warm water, free soap, free hand sanitizer, and free cleaning products, increased time spent outside, increased physical/social distancing measures, decreased population density achieved by releasing people, increased access to free medical care, and improved nutrition are all factors resulting in instant and obvious improvements in individual health outcomes for people incarcerated within the jail system. Alterations to function and practice of the jail system that can correct for these challenges are unlikely to occur quickly enough or substantially enough to improve the epidemiological risks for the incarcerated people within the jail system. As our results have shown, even when the within-jail transmission rates are improved by interventions such as reduction in intake from new arrests leading to a decrease in the size of the incarcerated population, we cannot effectively reduce the outbreak of infection in either the staff or incarcerated people down to the levels of the broader community.

As with all models, the conclusions of this study depend on an accurate representation of the flow of individuals between the jail system and the wider community, either due to arrests or due to their employment as jail staff, as well as the values of the parameters used to determine how swiftly this flow occurs. The inherent nature of emerging epidemics makes both of these things uncertain — the clinical and biological aspects of the pathogen might not be fully understood, and the data needed to parameterize these models is often sparse and incomplete. This problem is especially acute in models of this sort, which seek to present a “what-if” scenario to stave off a public health crisis, rather than analyze how that crisis unfolded after the fact. Nevertheless, while the exact projected magnitudes may be sensitive to these unknowns, in truth, the greatest utility of models such as these in in determining best courses of action and likely magnitudes of the effects that can be gained from those actions, rather than exact predictions of precise numbers of individuals (*31*). Due to the logical nature of the processes studied, so long as errors in the parameters used are consistent across scenarios, they will not impact the understanding that results from our projections about which courses of action achieve the best outcomes, even if those errors would alter our understanding of the precise amount of effect achieved by each intervention.

## Conclusion

Conditions within jails must be immediately improved to decrease the probabilities of disease transmission and support better health for incarcerated people to protect not only themselves, but also jail staff and the community at large. These recommendations are time-critical as the next wave of the pandemic threatens to overwhelm community healthcare access, meaning any action that can reduce burden should be considered a critical public good. Decreasing the incarcerated population density both directly decreases disease exposure, interrupting transmission dynamics, and also facilitates many other interventions. It is a natural result of reduced intake. We can achieve/enable many desired benefits with just that one, simple action, but to achieve maximal benefits to society, preventing the greatest burden from disease both within the jails and without, broad actions that include alterations to both intake and release and also many of the within-jail strategies for improving the individual means to enact personal hygiene, protection through social distancing, and access to medical care are all needed.

## Model/Methods

### Transmission Model

We begin by tailoring a standard SEIR model to the specific dynamics of COVID-19. We first split our total population into four categories of risk: Children under 18 (denoted with the subscript *K*), Low-risk adults (denoted with the subscript *L*), High-risk adults (denoted with the subscript *H*), and Elderly adults (denoted with the subscript *E*). We also designate a separate population category for jail staff, *O* (note: while *O* was the selected notation, it is meant to capture all staff working at the jail, not only the corrections officers). These populations are then assigned into disease-related health status compartments: Susceptible (*S*), Exposed (in which individuals are presymptomatic, but do already produce low levels of infection transmission to others, *E*), Infected (in which individuals are both symptomatic themselves and fully infectious to others, *I*), Medically Treated (those infectious individuals whose disease severity and healthcare access results in removal from the population into a medical care facility that prevents any further transmission of infection back into the population, *M*), and Removed (those who have either recovered from the infection and are now immune or those who have died, *R*). We also allow for the possibility that an Infected person with sufficient disease severity to warrant medical treatment is unable to obtain care, and designate rates associated with this case, as designated by the subscript *U*. For clarity of the results, we do not consider death from any non-COVID-19 cause; this is done to highlight the COVID-19-specific dynamics. Additionally, as a simplifying assumption due to their low rates of both infections and complications, we do not model hospitalizations or deaths in children. Similarly, once hospitalized, patients are assumed not to spread COVID-19 further, as additionally modeling the impact of healthcare-associated COVID-19 cases is well beyond the scope of this model. Lastly, we split our population into segments depending on the subsection of the community or jail system in which they are currently functioning: the community at large, *C*, the processing system for the jail, *P*, the court system *T*, and the jail system, *J*.

A schematic for the whole model can be seen in Fig. 7, and the differential equations comprising the model are in Supplementary Materials Appendix 1. Fig. 8 shows separately the model for transitions between locations (left) and the model for transitions between disease states (right). In this figure, we give the model parameters used to describe the rate of transition between locations and states. The model was implemented in R 3.6.3 using the de-Solve package, with the visualization of results primarily using ggplot2. Statistical analysis of one parameter (see below) was done using the flexsurv package. The code and data used in this analysis is available at https://www.github.com/epimodels/COVID19-Jails. As this study used only publicly available data and does not involve human subjects, IRB approval was not required.

**Fig. 7.**
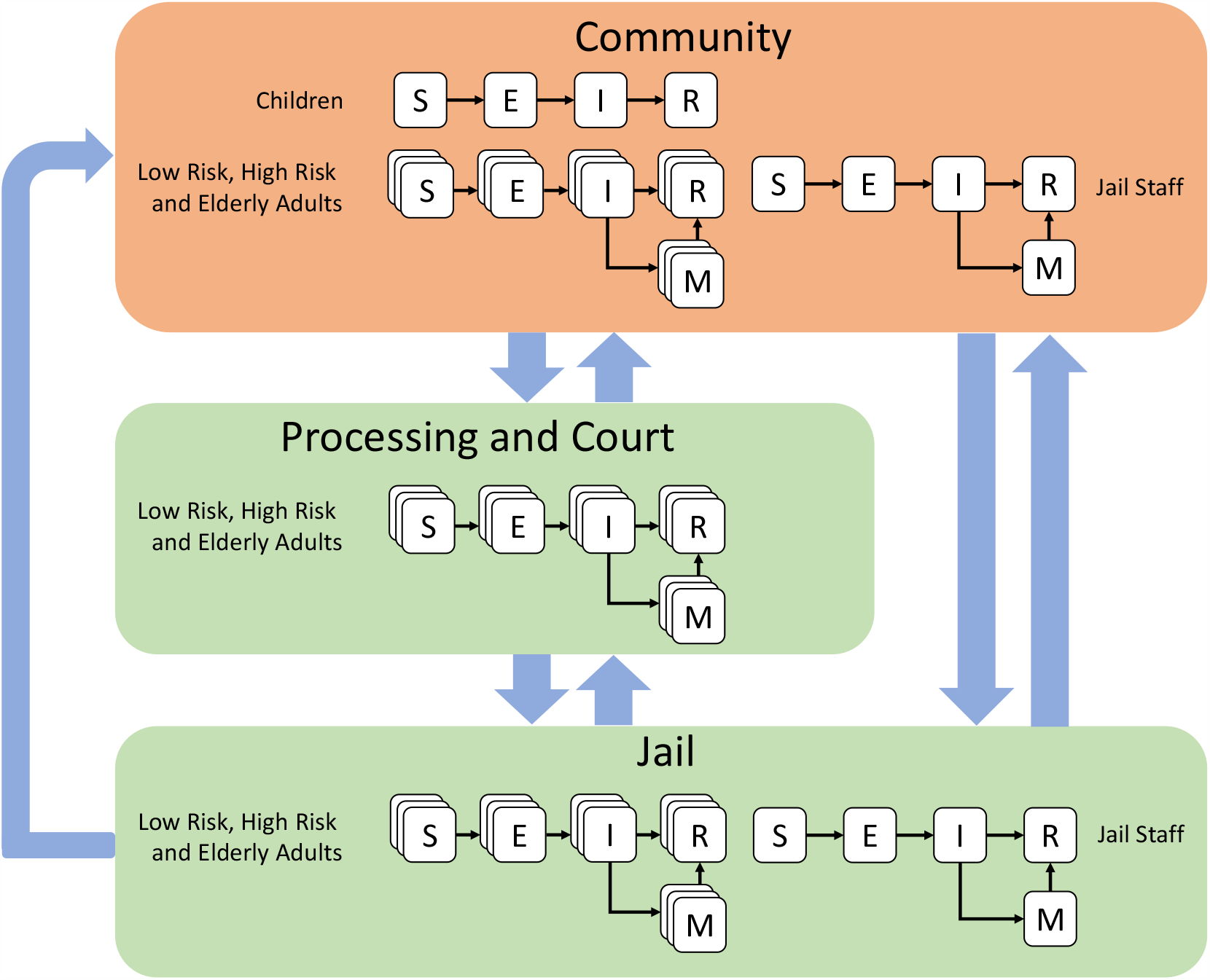
Schematic for a mathematical model of COVID-19 in a linked urban community - jail system. The population is represented in one of five possible compartments: Susceptible (S), Exposed (E), Infected (I), Needing Medical Care (M) and Recovered/Removed (R). Additionally, the population is divided into five distinct sub-populations: Children under 18 years of age, Elderly Adults over 65 years of age, Low Risk Adults between 18 and 65, High Risk Adults between 18 and 65 and Jail Staff (assumed to be between 18 and 65 years of age). Arrested adults move between the Community, Processing and the Court System and Jail, while Jail Staff move between the Community and Jail. Children are assumed not to be eligible for arrest.

**Fig. 8.**
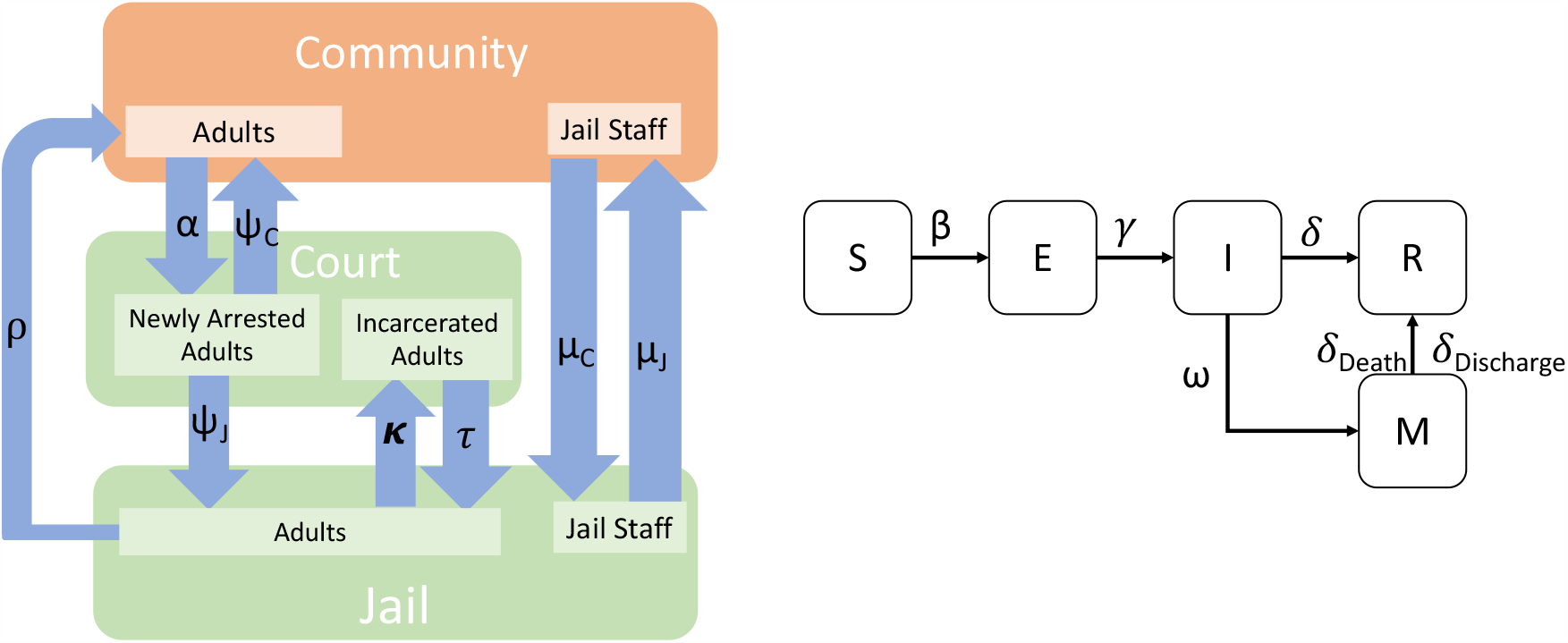
(left) Depiction of model emphasizing movement between locations. Here “Court” stands for both processing and trials. (right) Depiction of the model emphasizing transition between disease states. The parameter family is shown for each type of transition. For example, although we show just one *α* between community and court, in our model there are separate sub-scripted *α* parameters corresponding to different rates of arrest for different age groups. Similar logic applies to the other parameters.

### Population Movement Into, Within, and Out Of the Jail System

Our model captures movement between the community (denoted with superscript, *C*), processing (*P*),jail (denoted with superscript, *J*), and court appointments (denoted with *T* for trial, though this is meant to encompass all court appointments). It assumes staff move only between the community and jail; they are not arrested in our model. We base the parameters of movement into, within, and out of the jail on Allegheny County, Pennsylvania, where detailed data on the jail population and facilities are available, including an automatically updating dashboard giving statistics for the jail population (*32*). In Allegheny County, the population at large is approximately 1.2 million people. The size of the jail population hovers around 2,500. We use these population figures to initialize our model.

In our model, individuals in the community are arrested at a rate of approximately 100 people per day (*33*). Based on discussion with experts, arrest rates were calibrated such that approximately 40% of those arrested were at high risk for COVID-19, though the underlying conditions this represents may be very different from those prevalent in the community at large (e.g. high levels of immune suppression from drug use). Arrested individuals are brought to processing. From processing, individuals can either be released back into the community (60%) or taken to jail (40% (*32*)). This results in an in-flow to the jail of approximately 40 individuals per day, which is consistent with Allegheny County’s reporting. While in jail, individuals transition back and forth between the jail and court appointments. The number of movements between jail and court appointments is described as “well over 100” on the jail’s website. We assume movement of approximately 150 people per day between the jail and court. For each court appointment, we assume individuals spend half a day on average at the court facility. Importantly, we also assume that there is mixing in the court facility between those who are there for processing after arrest and those that are present there for court appointments. From the jail, individuals are released back into the community at a rate that is consistent with the reported 62 day average length of stay. One limitation of our model is that we do not account for post-jail destinations that are not the community, i.e. we do not model people moving from jail to prison. According to (*34*), the yearly number of admissions to prison is about 600,000 while the yearly number of admissions to jail is around 10.6 million. So, assuming that all prison admissions first had one jail admissions, around 95% of all jail admissions do not go on to prison; they are released back into the community as in our model. Thus, we expect that the omission of prison from our model does not substantially impact the overall findings.

An online database of public employees salaries in Allegheny County shows a population of 384 people whose job title is corrections officer, whose job location is the jail, and who are listed as active. Although this is certainly an underestimate of the total number of the jail’s staff, which includes other types of employees, we think this is a useful approximation to the total number of staff. We use this figure as the number of staff members moving between community and jail. Staff transition between community and jail at a rate that assumes 8 hour shift lengths in the jail per day with the remaining 16 hours per day spent in the community.

### Estimation Population Mixing and Contact Rates

To estimate the *β* parameters governing movement between disease states, we break the problem into two parts: estimating an unscaled matrix of transmission rates between age groups and calculating appropriate scaling factors. In summary, we use the methodology in (*35*) to create the matrix of the relative rate of transmission between age groups. Because each location (community, jail, processing/trial) is composed of different numbers of people, this matrix must be re-scaled by the population size in each location so that the mixing parameters have comparable meanings. We further scale according to the assumption that people in jail, processing, and trial mix at a rate that is approximately three, six, and six times that of people in the community, respectively. This is based on qualitative evidence of increased mixing rates in those locations. Finally, we select a common scale parameter, *c*_0_, across all locations that calibrates our model. Below we go into detail for each of the steps of this calculation.

We first estimate parameters that describe the relative rate of transmission between each of the community categories: Children, Low-risk adults, High-risk adults, and Elderly adults, from other studies. We denote the rate of transmission to category *q* from category *r* in the community by 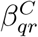. We decompose 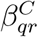 into a scale parameter *β*^*C*^ and matrix of scale-free *relative* transmission rates, 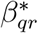.

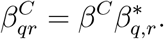

To estimate the relative transmission rates, 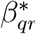, we follow the methodology outlined in (*35*) for modeling the spread of COVID-19 in the United States. As in (*35*), we define 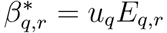, where *u*_*q*_ is the susceptibility of people in compartment *q, E*_*q,r*_ are the entries of the contact matrix denoting the mean number of contacts between individuals in the *q*th and *r*th compartments. As in (*35*), we use consensus estimates of susceptibility from (*36*). Our model has fewer age groups than reported in (*36*). We thus define *u*_*q*_ as a population-weighted average of the values reported in (*36*). For example, in our model we define the children compartment to contain people aged 0-17. (*36*) reports *u*_0*−*9_ and *u*_10*−*19_, the average susceptibility of individuals in the 0-9 and 10-19 age groups, respectively. To create our *u*_*K*_, we set 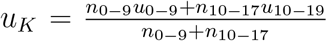 where *n*_0*−*9_ is the number of people in Allegheny County aged 0-9 and *n*_10*−*17_ is the number of people in Allegheny County aged 10-17. We obtain population totals for Allegheny County from the US Census. We use the socialmixr package R package (*37*) to obtain the values of *E*_*q,r*_. As in (*35*), we use the contact matrix for the United Kingdom as a stand-in for contact rates in the United States. The resulting values of 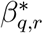 is shown in Table 1.

**Table 1:** Relative transmission rates of COVID-19 from an individual per row to an individual per column.

|  | child | adult | elderly |
| --- | --- | --- | --- |
| child | 2.72 | 1.60 | 0.93 |
| adult | 1.92 | 6.22 | 4.39 |
| elderly | 0.09 | 0.49 | 1.38 |

For each of the non-community locations, we assume that mixing patterns are not category-dependent. That is, we assume that all incarcerated people mix equally, regardless of age. Mathematically, this amounts to the assumption that 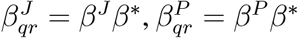, and 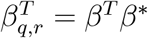, where *β*^*\**^ is defined to be consistent with the *β*^*\**^s in the community, and *β*^*J*^, *β*^*P*^, and *β*^*T*^ are scaling parameters. Because most of the people in jail, processing, and trial are adults, we take 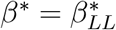.

This leaves setting the scale parameters, *β*^*C*^, *β*^*J*^, *β*^*P*^, and *β*^*T*^. There are two considerations to account for here: differences in the size of the population in each of the four locations, and differences in contact rates within the four locations. We define 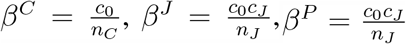, and 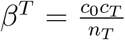. Where *n*_*c*_ = 1, 200, 000, *n*_*j*_ = 2, 600, *n*_*t*_ = 150, and *n*_*p*_ = 100 are the size of the population in the community, jail, and processing, respectively. Scaling each of the *β* terms in the model by the population sizes amounts to the assumption that transmission is contact-based. This assumption leads to conservative estimates of the speed of the spread in the jail system relative to a fomite-based transmission model. The parameters *c*_*j*_, *c*_*p*_, and *c*_*t*_ are factors that denotes how many times more contacts per day a person in jail or processing, respectively, has than a person in the community. We set these values to be *c*_*j*_ = 3 and *c*_*p*_ = *c*_*t*_ = 6, corresponding to an assumption of three and six times more contact in jail and processing/trial, respectively, than take place in the community. These values were chosen to reflect the conditions for both health and crowding of populations within jail facilities (*21–26*), and the understood gathering and transportation protocols associated with intake*/*release processing and court appearances.

The sensitivity of the model to these and other parameter choices is explored in SI Appendix 2. An analysis of the sensitivity of the model to perturbations of the parameter values allows us to determine how changes in parameters (due to variation over time, uncertainty in measurement, or otherwise) impacts the quantitative outcomes of the model. (*38*) It is possible to use such a sensitivity analysis to identify the more influential parameters, e.g. (*39*).

To calibrate the model for our baseline scenario, we then find a *c*_0_ such that approximately 80% of the population is ultimately infected by the time the spread dies out in our model. We selected an 80% final infection rate for consistency with predictions of the spread of COVID-19 under the assumption of no mitigation measures in place from an influential micro-simulation model (*40*). To calibrate the model for our shelter-in-place scenario, we select a *c*_0_ that matches the trajectory of COVID-19 in Allegheny County under shelter-in-place. Specifically, we calibrate our model to the time series of deaths in Allegheny county as reported in data released by the New York Times. Because very few infections or deaths were reported prior to the initial shelter-in-place order on March 23, 2020, we assume the beginning of the death time series all took place under shelter-in-place conditions. On May 15, the county entered the “yellow phase” in which businesses began re-opening. We assume that it takes approximately a month for infections that occurred under shelter-in-place to result in death, so we take the total number of deaths that occurred under shelter-in-place to be the total number of deaths as of June 15, 2020. There were 174 deaths that occurred as of this date. We set the start-date of the epidemic to be one month prior to the first death, February 20, 2020. We then perform a grid search to find a shelter-in-place scalar for *c*_0_ such that over the course of approximately 115 days between February 20 and Jun 15, approximately 174 deaths occurred in our simulation and the trajectories are similar. Fig. 9 shows a comparison between the time series of COVID-19 deaths in Allegheny County and the trajectory produced by the *c*_0_ scalar that corresponds to the best fit.

**Fig. 9.**
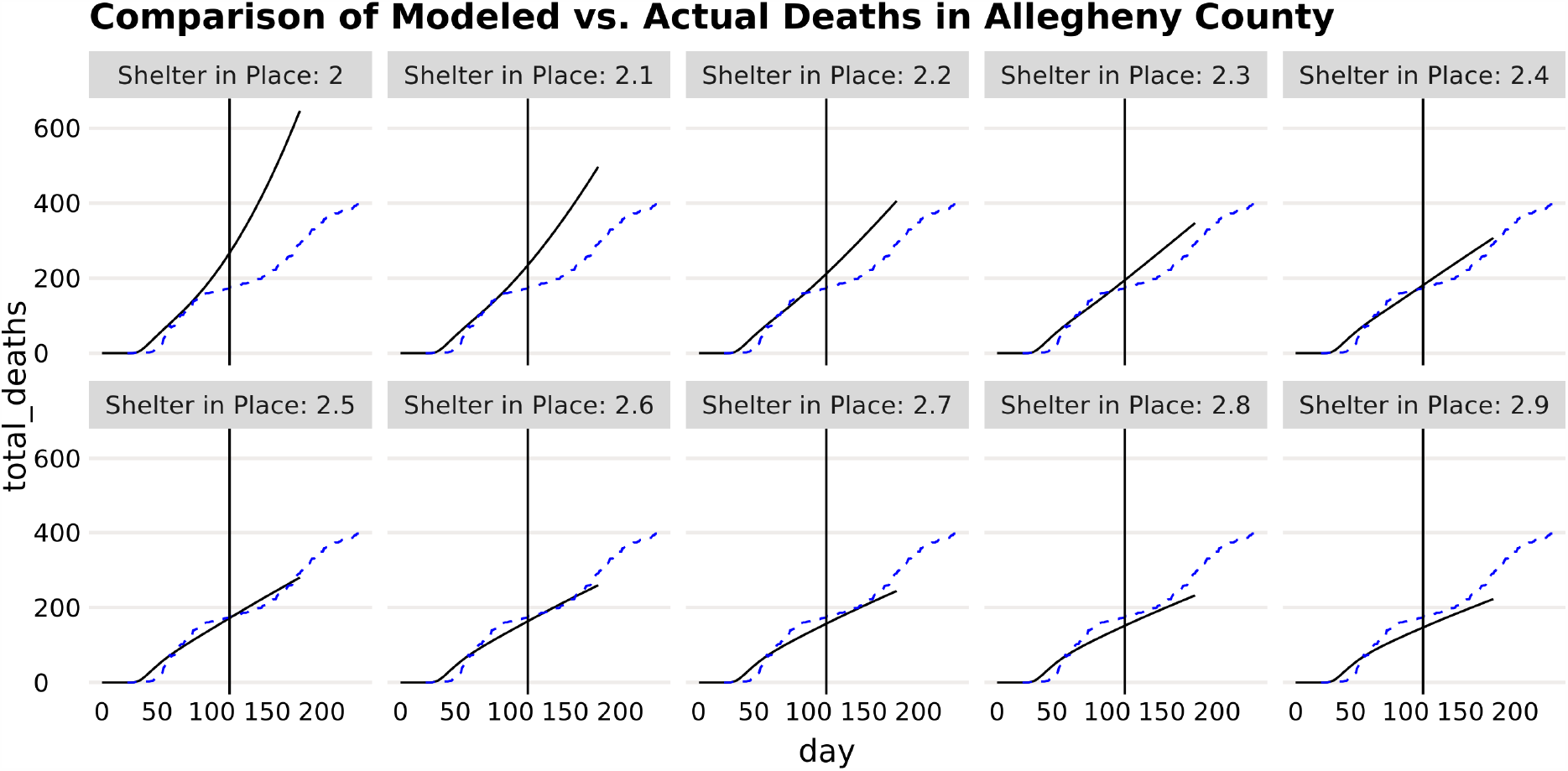
Actual vs. modelled deaths under a grid search of various shelter-in-place scalar coefficients. Each graph divides the baseline *c*_0_ by the Shelter-in-Place scalar shown in the graph legends, effectively reducing the *β* coefficients from baseline. Here, the factor with the best fit was 2.5. The vertical line shows June 15, 30 days after Allegheny relaxed social distancing.

### Estimation of Other Model Parameters

Parameters concerning the natural history of COVID-19, patient progression, etc. were primarily obtained from existing estimates in the modeling literature, where possible using estimates from as close to the modeled catchment area as possible (i.e. CHIME from UPenn Medicine, https://penn-chime.phl.io/). Citations for specific parameter values may be found in Table 2.

**Table 2:**
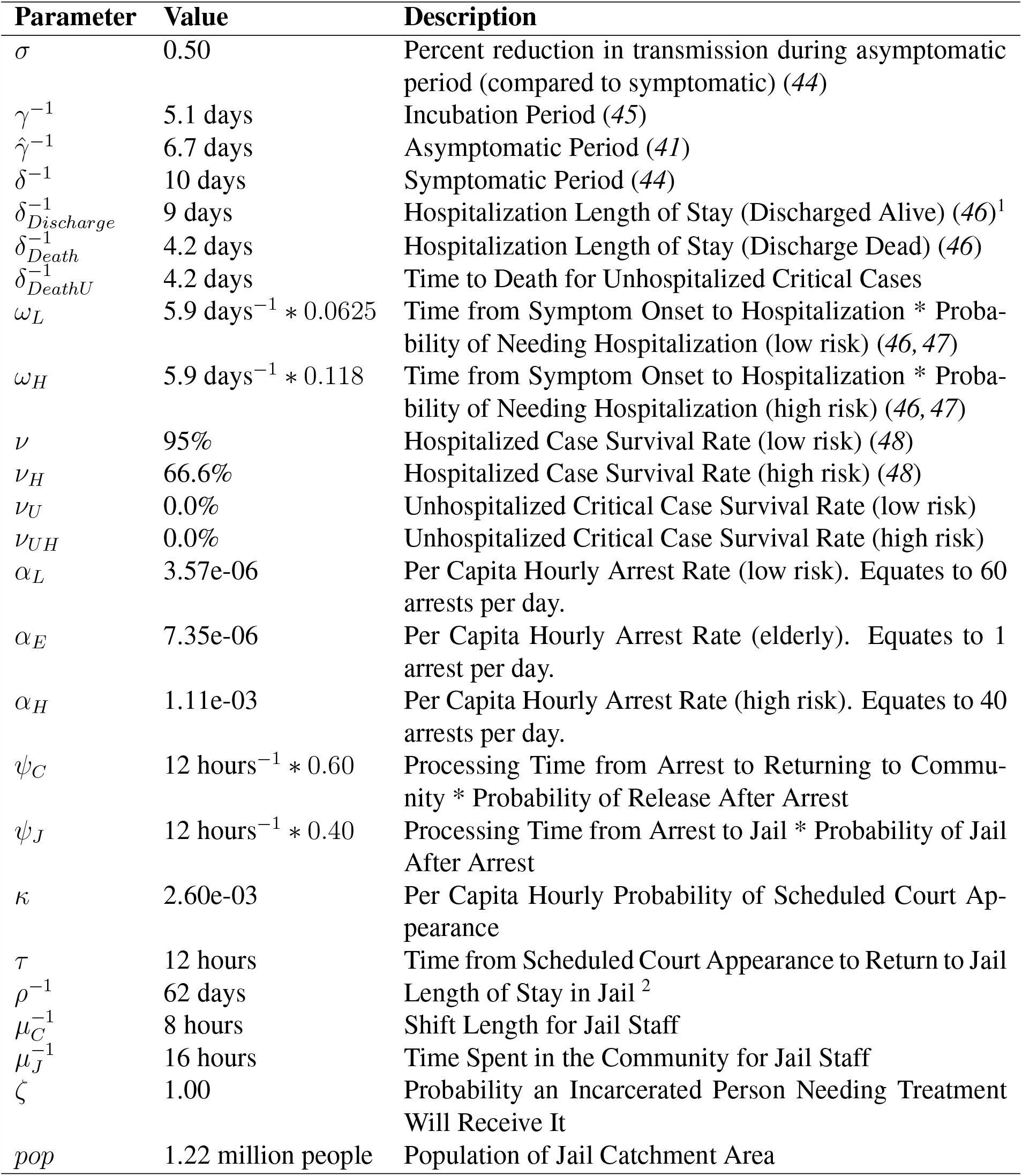
Parameter values, meanings and sources for a community-jail transmission model of COVID-19.

| Parameter | Value | Description |
| --- | --- | --- |
| $\sigma$ | 0.50 | Percent reduction in transmission during asymptomatic period (compared to symptomatic) (44) |
| $\gamma^{-1}$ | 5.1 days | Incubation Period (45) |
| $\hat{\gamma}^{-1}$ | 6.7 days | Asymptomatic Period (41) |
| $\delta^{-1}$ | 10 days | Symptomatic Period (44) |
| $\delta_{Discharge}^{-1}$ | 9 days | Hospitalization Length of Stay (Discharged Alive) (46)<sup>1</sup> |
| $\delta_{Death}^{-1}$ | 4.2 days | Hospitalization Length of Stay (Discharge Dead) (46) |
| $\delta_{DeathU}^{-1}$ | 4.2 days | Time to Death for Unhospitalized Critical Cases |
| $\omega_L$ | 5.9 days<sup>-1</sup> * 0.0625 | Time from Symptom Onset to Hospitalization * Probability of Needing Hospitalization (low risk) (46, 47) |
| $\omega_H$ | 5.9 days<sup>-1</sup> * 0.118 | Time from Symptom Onset to Hospitalization * Probability of Needing Hospitalization (high risk) (46, 47) |
| $\nu$ | 95% | Hospitalized Case Survival Rate (low risk) (48) |
| $\nu_H$ | 66.6% | Hospitalized Case Survival Rate (high risk) (48) |
| $\nu_U$ | 0.0% | Unhospitalized Critical Case Survival Rate (low risk) |
| $\nu_{UH}$ | 0.0% | Unhospitalized Critical Case Survival Rate (high risk) |
| $\alpha_L$ | 3.57e-06 | Per Capita Hourly Arrest Rate (low risk). Equates to 60 arrests per day. |
| $\alpha_E$ | 7.35e-06 | Per Capita Hourly Arrest Rate (elderly). Equates to 1 arrest per day. |
| $\alpha_H$ | 1.11e-03 | Per Capita Hourly Arrest Rate (high risk). Equates to 40 arrests per day. |
| $\psi_C$ | 12 hours<sup>-1</sup> * 0.60 | Processing Time from Arrest to Returning to Community * Probability of Release After Arrest |
| $\psi_J$ | 12 hours<sup>-1</sup> * 0.40 | Processing Time from Arrest to Jail * Probability of Jail After Arrest |
| $\kappa$ | 2.60e-03 | Per Capita Hourly Probability of Scheduled Court Appearance |
| $\tau$ | 12 hours | Time from Scheduled Court Appearance to Return to Jail |
| $\rho^{-1}$ | 62 days | Length of Stay in Jail <sup>2</sup> |
| $\mu_C^{-1}$ | 8 hours | Shift Length for Jail Staff |
| $\mu_J^{-1}$ | 16 hours | Time Spent in the Community for Jail Staff |
| $\zeta$ | 1.00 | Probability an Incarcerated Person Needing Treatment Will Receive It |
| $pop$ | 1.22 million people | Population of Jail Catchment Area |

In one case, 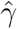, or the asymptomatic period^*−*1^, the original source reported that their estimate was likely an underestimation due to censoring. However, given that the authors provided the data within their manuscript (*41*), the data was re-estimated to account for censoring using a parametric survival model assuming an exponential distribution (the distribution typically implied by the uniform hazard of transitioning from one compartment to another within a compartmental model). The fit for this exponential model may be found in SM Appendix 3.

### Modeled Scenarios and Interventions

We represented the effects of several policy interventions or failures as changes to various parameters in this model. We consider four categories of scenarios that could vary the rate of spread: in addition to modeling shelter-in-place (reduced mixing) conditions in the community, we modeled scenarios related to reductions in arrest rates, increases in release rates, and changes to within-jail conditions. These scenarios are detailed below in Table 3. Most scenarios are additive; that is, all arrest reduction interventions assume a baseline scenario of shelter-in-place in the community. The scenarios involving faster release of individuals in jail all assume both shelter-in-place in the community, and were each run under each of the “Arrest Reduction” scenarios to determine the cumulative effects of arrest reduction, increased release rates, and community shelter-in-place conditions. The mixing reduction scenario in the jail assumes shelter-in-place in the community as well as a 25% reduction in arrests (equivalent to the “Bail Eligible” Arrest Reduction scenario), as it is unlikely that jails will be able to effectively reduce contact rates without reducing their average daily population. Finally, in the reduced detection scenario, we assume shelter-in-place, but vary the likelihood that serious cases of COVID-19 is caught and treated in a timely manner.

**Table 3:** Scenarios and parameter adjustments for a number of policy-based interventions to curtail COVID-19 in jail and the community.

| Scenario Name | Parameters | Multiplier of Baseline | Scenario Description |
| --- | --- | --- | --- |
| Shelter in Place | $\beta_*^C$ | 0.4 | Effective contact rate in the community is reduced by a factor of 1/2.5. |
| *Arrest Reduction* |  |  |  |
| Bail Eligible | $\alpha_L, \alpha_H, \alpha_E$ | 0.75 | Divert all bail-eligible arrests (estimated at 25% of all arrests) |
| Vulnerable Only | $\alpha_H, \alpha_E$ | 0.10 | Divert arrests of 90% of vulnerable populations. (11) |
| Low Level | $\alpha_L, \alpha_H, \alpha_E$ | 0.17 | Divert all low-level arrests (estimated at 83% of arrests). |
| Arrest Fewer People | $\alpha_L, \alpha_H, \alpha_E$ | 0.10 | Divert 90% of current arrests. |
| *Faster Release* |  |  |  |
| Increase Release Speed | $\rho^{-1}$ | 2 | 2x rate of release from jail |
| Vulnerable Only | $\rho_H^{-1}$ | 2 | 2x rate of release for vulnerable only |
| *In-Jail Scenarios* |  |  |  |
| Mixing Reduction | $\beta_T, \beta_P, \beta_J$ | 0.4 | Reduction of baseline contact rates in jails by the same factor as the community under shelter-in-place |
| Reduced Detection | $\zeta$ | 0.99,0.95,0.90 | Reduction in infection detection and timely hospitalization in jails by 1-ζ |

Below, vulnerable populations are defined as individuals over the age of 65 or at increased risk of complications form COVID-19 due to other co-morbidities. We estimate that 40% of the jail population is vulnerable by this definition, based on reported rates of co-morbidities among incarcerated people (*42*). We estimate that around 25% of those arrested are bail eligible, based on information from Allegheny county on the rate at which cash bail was used between February and June of 2019 (*43*).

## Data Availability

The code necessary to run the models is available upon request from the authors, and the equations and parameter values necessary for the results of the manuscript are contained within it.

## Acknowledgments

EL was supported by the CDC Cooperative Agreement RFA-CK-17-001-Modeling Infectious Diseases in Healthcare Program (MInD-Healthcare). *Author Contributions:* EL, KL, and AH conceived of the study. EL, KL, and NF designed the model. EL, KL, AH, BM, and NF all contributed to parameter estimation. EL, KL, and AH implemented the model. AH and BM designed the figures. KM performed the sensitivity analyses. All authors contributed to the interpretation of the results and preparation of the manuscript. *Competing Interest:* AH and BM are employed as researchers at an advocacy organization, in collaboration with coauthors who are unpaid members of that same advocacy organization (EL, KL, KM, NF). That organization had no role in determining how the study was conducted, or the results obtained. Further, EL and NF are currently acting as pro bono expert witnesses for legal cases based on the results of this work. *Data and Material Availability:* All data used was gathered from publicly available, published sources, as referenced in the text and tables.

## References

1. S. Mervosh, D. Lu, V. Swales, New York Times (2020).

2. J. Bayham, E. P. Fenichel, Available at SSRN 3555259 (2020).

3. W. E. Parmet, M. S. Sinha, New England Journal of Medicine 382, e28 (2020).

4. H. Bernstein, J. González, D. Gonzalez, J. Jagannath (2020).

5. R. Sharp (2018).

6. B. L. Sykes, B. Pettit, Handbook on Children with Incarcerated Parents (Springer, 2019), pp. 11–23.

7. R. R. Weidner, J. Schultz, SSM-Population Health 9, 100466 (2019).

8. S. Kelley, Human Ecology 47, 15 (2019).

9. P. K. Enns, et al., Socius 5, 2378023119829332 (2019).

10. H. D. Lester, M. J. Miller, Computer Security (Springer, 2019), pp. 307–322.

11. T. D. Minton, Bureau of Justice Statistics (2011).

12. S. Raher, The company store: A deeper look at prison commissaries (2018).

13. J. A. Bick, Clinical Infectious Diseases 45, 1047 (2007).

14. C. W. Hoge, et al., New England Journal of Medicine 331, 643 (1994). PMID: 8052273.

15. R. Rubin, Jama 323, 1760 (2020).

16. E. Reinhart, D. Chen, Health Affairs pp. 10–1377 (2020).

17. M. J. Akiyama, A. C. Spaulding, J. D. Rich, New England Journal of Medicine 382, 2075 (2020).

18. L. Hawks, S. Woolhandler, D. McCormick, JAMA Internal Medicine (2020).

19. M. Thigpen, T. Beauclair, V. Hutchinson, V. Persons, C. Paul, Jail design guide (2011).

20. C. for Disease Control, Prevention, et al., Interim guidance on management of coronavirus disease 2019 (covid-19) in correctional and detention facilities (2020).

21. K. M. Nowotny, Health & justice 5, 9 (2017).

22. S. Mignon, Ciencia & saude coletiva 21, 2051 (2016).

23. E. S. Barnert, R. Perry, R. E. Morris, Academic pediatrics 16, 99 (2016).

24. D. C. McClelland, C. Alexander, E. Marks, Journal of Abnormal Psychology 91, 61 (1982).

25. E. T. Jacobs, C. J. Mullany, Nutrition 31, 659 (2015).

26. F. G. Kouyoumdjian, E. M. Andreev, R. Borschmann, S. A. Kinner, A. McConnon, PloS one 12 (2017).

27. Allegheny county jail population management: Interactive dashboards (2020).

28. I. A. Binswanger, P. J. Blatchford, S. J. Forsyth, M. F. Stern, S. A. Kinner, Public Health Reports 131, 574 (2016).

29. T. Williams, L. Seline, R. Griesbach, New York Times. June 18 (2020).

30. J. Ransom, New York Times. May 20 (2020).

31. E. T. Lofgren, et al., Proceedings of the National Academy of Sciences 111, 18095 (2014).

32. Allegheny County Analytics. https://perma.cc/93RG-4WZ8.

33. Allegheny County Analytics. https://perma.cc/9DSP-9CTY.

34. P. Wagner, L. Sakala, Prison Policy Initiative 12 (2014).

35. I. F. Miller, A. D. Becker, B. T. Grenfell, C. J. E. Metcalf, Nature Medicine 26, 1212 (2020).

36. N. G. Davies, et al., MedRxiv (2020).

37. . S. Funk (2018).

38. J. Wu, R. Dhingra, M. Gambhir, J. V. Remais, Journal of The Royal Society Interface 10, 20121018 (2013).

39. A. Lurette, S. Touzeau, M. Lamboni, H. Monod, Journal of Theoretical Biology 258, 43 (2009).

40. N. Ferguson, et al. (2020).

41. Z. Hu, et al., Science China Life Sciences pp. 1–6 (2020).

42. L. M. Maruschak, M. Berzofsky, J. Unangst, Medical problems of state and federal prisoners and jail inmates, 2011-12 (US Department of Justice, Office of Justice Programs, Bureau of Justice …, 2015).

43. A. Pennsylvania, Punishing poverty: Cash bail in allegheny county (2019).

44. J. S. Weitz, Available on GitHub (2020).

45. S. A. Lauer, et al., Annals of internal medicine (2020).

46. R. Verity, et al., MedRxiv (2020).

47. L. Tindale, et al., medRxiv (2020).

48. C. COVID.

49. M. Stein, Technometrics 29, 143 (1987).

