## Supplementary Information for "The Epidemiological Implications of Jails for Community, Corrections Officer, and Incarcerated Population Risks from COVID-19"

### Supplementary Materials

**SM Appendix 1.** According to the logic presented in the Model/Methods section 7 define the following system of equations to capture the epidemiological dynamics of our system:

Within the Broader Community:

$$\begin{aligned}\frac{dS_K^C}{dt} &= -\beta_{KK}^C S_K^C \left(I_K^C + \sigma E_K^C\right) - \beta_{KL}^C S_K^C \left(I_L^C + I_O^C + \sigma\left(E_L^C + E_O^C\right)\right)\\ &\quad - \beta_{KE}^C S_K^C \left(I_E^C + \sigma E_E^C\right) - \beta_{KH}^C S_K^C \left(I_H^C + \sigma E_H^C\right)\end{aligned}$$

$$\begin{aligned}\frac{dE_K^C}{dt} &= \beta_{KK}^C S_K^C \left(I_K^C + \sigma E_K^C\right) + \beta_{KL}^C S_K^C \left(I_L^C + I_O^C + \sigma\left(E_L^C + E_O^C\right)\right)\\ &\quad + \beta_{KE}^C S_K^C \left(I_E^C + \sigma E_E^C\right) + \beta_{KH}^C S_K^C \left(I_H^C + \sigma E_H^C\right) - \left(\gamma + \hat{\gamma}\right) E_K^C\end{aligned}$$

$$\frac{dI_K^C}{dt} = \gamma E_K^C - \delta I_K^C$$

$$\frac{dR_K^C}{dt} = \delta I_K^C + \hat{\gamma} E_K^C$$

$$\begin{aligned}\frac{dS_L^C}{dt} &= -\beta_{LK}^C S_L^C \left(I_K^C + \sigma E_K^C\right) - \beta_{LL}^C S_L^C \left(I_L^C + I_O^C + \sigma\left(E_L^C + E_O^C\right)\right)\\ &\quad - \beta_{LE}^C S_L^C \left(I_E^C + \sigma E_E^C\right) - \beta_{LH}^C S_L^C \left(I_H^C + \sigma E_H^C\right) - \alpha_L S_L^C + \psi_C S_L^P + \rho S_L^J\end{aligned}$$

$$\begin{aligned}\frac{dE_L^C}{dt} &= \beta_{LK}^C S_L^C \left(I_K^C + \sigma E_K^C\right) + \beta_{LL}^C S_L^C \left(I_L^C + I_O^C + \sigma\left(E_L^C + E_O^C\right)\right)\\ &\quad + \beta_{LE}^C S_L^C \left(I_E^C + \sigma E_E^C\right) + \beta_{LH}^C S_L^C \left(I_H^C + \sigma E_H^C\right) - \left(\gamma + \hat{\gamma}\right) E_L^C\\ &\quad - \alpha_L E_L^C + \psi_C E_L^P + \rho E_L^J\end{aligned}$$

$$\frac{dI_L^C}{dt} = \gamma E_L^C - \delta I_L^C - \omega_L I_L^C - \alpha_L I_L^C + \psi_C I_L^P + \rho I_L^J$$

$$\frac{dM_L^C}{dt} = \omega_L I_L^C - \delta_{Discharge} M_L^C - \delta_{Death} \nu M_L^C$$

$$\begin{aligned}\frac{dR_L^C}{dt} &= \delta I_L^C + \hat{\gamma} E_L^C + \delta_{Discharge} M_L^C + \delta_{Death} \nu M_L^C - \alpha_L R_L^C + \psi_C R_L^P\\ &\quad + \rho R_L^J\end{aligned}$$

$$\begin{aligned}\frac{dS_E^C}{dt} &= -\beta_{EK}^C S_E^C \left(I_K^C + \sigma E_K^C\right) - \beta_{EL}^C S_E^C \left(I_L^C + I_O^C + \sigma\left(E_L^C + E_O^C\right)\right)\\ &\quad - \beta_{EE}^C S_E^C \left(I_E^C + \sigma E_E^C\right) - \beta_{EH}^C S_E^C \left(I_H^C + \sigma E_H^C\right) - \alpha_E S_E^C + \psi_C S_E^P + \rho S_E^J\end{aligned}$$

$$\frac{dE_E^C}{dt} = \beta_{EK}^C S_E^C \left(I_K^C + \sigma E_K^C\right) + \beta_{EL}^C S_E^C \left(I_L^C + I_O^C + \sigma\left(E_L^C + E_O^C\right)\right) + \beta_{EE}^C S_E^C \left(I_E^C + \sigma E_E^C\right) + \beta_{EH}^C S_E^C \left(I_H^C + \sigma E_H^C\right) - (\gamma + \hat{\gamma}) E_E^C - \alpha_E E_E^C + \psi_C E_E^P + \rho E_E^J$$

$$\frac{dI_E^C}{dt} = \gamma E_E^C - \delta I_E^C - \omega_H I_E^C - \alpha_E I_E^C + \psi_C I_E^P + \rho I_E^J$$

$$\frac{dM_E^C}{dt} = \omega_H I_E^C - \delta_{Discharge} M_E^C - \delta_{Death} \nu_H M_E^C$$

$$\frac{dR_E^C}{dt} = \delta I_E^C + \hat{\gamma} E_E^C + \delta_{Discharge} M_E^C + \delta_{Death} \nu_H M_E^C - \alpha_H R_E^C + \psi_C R_E^P + \rho R_E^J$$

$$\frac{dS_H^C}{dt} = -\beta_{HK}^C S_H^C \left(I_K^C + \sigma E_K^C\right) - \beta_{HL}^C S_H^C \left(I_L^C + I_O^C + \sigma\left(E_L^C + E_O^C\right)\right) - \beta_{HE}^C S_H^C \left(I_E^C + \sigma E_E^C\right) - \beta_{HH}^C S_H^C \left(I_H^C + \sigma E_H^C\right) - \alpha_H S_H^C + \psi_C S_H^P + \rho S_H^J$$

$$\frac{dE_H^C}{dt} = \beta_{HK}^C S_H^C \left(I_K^C + \sigma E_K^C\right) + \beta_{HL}^C S_H^C \left(I_L^C + I_O^C + \sigma\left(E_L^C + E_O^C\right)\right) + \beta_{HE}^C S_H^C \left(I_E^C + \sigma E_E^C\right) + \beta_{HH}^C S_H^C \left(I_H^C + \sigma E_H^C\right) - (\gamma + \hat{\gamma}) E_H^C - \alpha_H E_H^C + \psi_C E_H^P + \rho E_H^J$$

$$\frac{dI_H^C}{dt} = \gamma E_H^C - \delta I_H^C - \omega_H I_H^C - \alpha_H I_H^C + \psi_C I_H^P + \rho I_H^J$$

$$\frac{dM_H^C}{dt} = \omega_H I_H^C - \delta_{Discharge} M_H^C - \delta_{Death} \nu_H M_H^C$$

$$\frac{dR_H^C}{dt} = \delta I_H^C + \hat{\gamma} E_H^C + \delta_{Discharge} M_H^C + \delta_{Death} \nu_H M_H^C - \alpha_H R_H^C + \psi_C R_H^P + \rho R_H^J$$

$$\frac{dS_O^C}{dt} = -\beta_{OK}^C S_O^C \left(I_K^C + \sigma E_K^C\right) - \beta_{OL}^C S_O^C \left(I_L^C + I_O^C + \sigma\left(E_L^C + E_O^C\right)\right) - \beta_{OE}^C S_O^C \left(I_E^C + \sigma E_E^C\right) - \beta_{OH}^C S_O^C \left(I_H^C + \sigma E_H^C\right) - \mu_J S_O^C + \mu_C S_O^J$$

$$\frac{dE_O^C}{dt} = \beta_{OK}^C S_O^C \left(I_K^C + \sigma E_K^C\right) + \beta_{OL}^C S_O^C \left(I_L^C + I_O^C + \sigma\left(E_L^C + E_O^C\right)\right) + \beta_{OE}^C S_O^C \left(I_E^C + \sigma E_E^C\right) + \beta_{OH}^C S_O^C \left(I_H^C + \sigma E_H^C\right) - (\gamma + \hat{\gamma}) E_O^C - \mu_J E_O^C + \mu_C E_O^J$$

$$\frac{dI_O^C}{dt} = \gamma E_O^C - \delta I_O^C - \omega_L I_O^C - \mu_J I_O^C + \mu_C I_O^J$$

$$\frac{dM_O^C}{dt} = \omega_L \left(I_O^C + I_O^J\right) - \delta_{Discharge} \nu M_O^C - \delta_{Death} (1 - \nu) M_O^C$$

$$\frac{dR_O^C}{dt} = \delta I_O^C + \hat{\gamma} E_O^C + \delta_{Discharge} \nu M_O^C + \delta_{Death}(1-\nu) M_O^C - \mu_J R_O^C + \mu_C R_O^J$$

Within the Processing System:

$$\frac{dS_L^P}{dt} = -\beta_{LL}^P S_L^P \left(I_L^P + I_L^T + \sigma\left(E_L^P + E_L^T\right)\right) - \beta_{LE}^P S_L^P \left(I_E^P + I_E^T + \sigma\left(E_E^P + E_E^T\right)\right)$$
$$- \beta_{LH}^P S_L^P \left(I_H^P + I_H^T + \sigma\left(E_H^P + E_H^T\right)\right) + \alpha_L S_L^C - \psi_C S_L^P - \psi_J S_L^P$$

$$\frac{dE_L^P}{dt} = \beta_{LL}^P S_L^P \left(I_L^P + I_L^T + \sigma\left(E_L^P + E_L^T\right)\right) + \beta_{LE}^P S_L^P \left(I_E^P + I_E^T + \sigma\left(E_E^P + E_E^T\right)\right)$$
$$+ \beta_{LH}^P S_L^P \left(I_H^P + I_H^T + \sigma\left(E_H^P + E_H^T\right)\right) - (\gamma + \hat{\gamma}) E_L^P + \alpha_L E_L^C$$
$$- \psi_C E_L^P - \psi_J E_L^P$$

$$\frac{dI_L^P}{dt} = \gamma E_L^P - \delta I_L^P + \alpha_L I_L^C - \psi_C I_L^P - \psi_J I_L^P$$

$$\frac{dR_L^P}{dt} = \alpha_L R_L^C - \psi_C R_L^P - \psi_J R_L^P + \delta I_L^P + \hat{\gamma} E_L^P$$

$$\frac{dS_E^P}{dt} = -\beta_{EL}^P S_E^P \left(I_L^P + I_L^T + \sigma\left(E_L^P + E_L^T\right)\right) - \beta_{EE}^P S_E^P \left(I_E^P + I_E^T + \sigma\left(E_E^P + E_E^T\right)\right)$$
$$- \beta_{EH}^P S_E^P \left(I_H^P + I_H^T + \sigma\left(E_H^P + E_H^T\right)\right) + \alpha_E S_E^C - \psi_C S_E^P - \psi_J S_E^P$$

$$\frac{dE_E^P}{dt} = \beta_{EL}^P S_E^P \left(I_L^P + I_L^T + \sigma\left(E_L^P + E_L^T\right)\right) + \beta_{EE}^P S_E^P \left(I_E^P + I_E^T + \sigma\left(E_E^P + E_E^T\right)\right)$$
$$+ \beta_{EH}^P S_E^P \left(I_H^P + I_H^T + \sigma\left(E_H^P + E_H^T\right)\right) - (\gamma + \hat{\gamma}) E_E^P + \alpha_E E_E^C$$
$$- \psi_C E_E^P - \psi_J E_E^P$$

$$\frac{dI_E^P}{dt} = \gamma E_E^P - \delta I_E^P + \alpha_E I_E^C - \psi_C I_E^P - \psi_J I_E^P$$

$$\frac{dR_E^P}{dt} = \alpha_E R_E^C - \psi_C R_E^P - \psi_J R_E^P + \delta I_E^P + \hat{\gamma} E_E^P$$

$$\frac{dS_H^P}{dt} = -\beta_{HL}^P S_H^P \left(I_L^P + I_L^T + \sigma\left(E_L^P + E_L^T\right)\right) - \beta_{HE}^P S_H^P \left(I_E^P + I_E^T + \sigma\left(E_E^P + E_E^T\right)\right)$$
$$- \beta_{HH}^P S_H^P \left(I_H^P + I_H^T + \sigma\left(E_H^P + E_H^T\right)\right) + \alpha_H S_H^C - \psi_C S_H^P - \psi_J S_H^P$$

$$\frac{dE_H^P}{dt} = \beta_{HL}^P S_H^P \left(I_L^P + I_L^T + \sigma\left(E_L^P + E_L^T\right)\right) + \beta_{HE}^P S_H^P \left(I_E^P + I_E^T + \sigma\left(E_E^P + E_E^T\right)\right)$$
$$+ \beta_{HH}^P S_H^P \left(I_H^P + I_H^T + \sigma\left(E_H^P + E_H^T\right)\right) - (\gamma + \hat{\gamma}) E_H^P + \alpha_H E_H^C$$
$$- \psi_C E_H^P - \psi_J E_H^P$$

$$\frac{dI_H^P}{dt} = \gamma E_H^P - \delta I_H^P + \alpha_H I_H^C - \psi_C I_H^P - \psi_J I_H^P$$

$$\frac{dR_H^P}{dt} = \alpha_H R_H^C - \psi_C R_H^P - \psi_J R_H^P + \delta I_H^P + \hat{\gamma} E_H^P$$

Within the Trial System:

$$\begin{aligned}
\frac{dS_L^T}{dt} &= -\beta_{LL}^T S_L^T \left(I_L^P + I_L^T + \sigma\left(E_L^P + E_L^T\right)\right) - \beta_{LE}^T S_L^T \left(I_E^P + I_E^T + \sigma\left(E_E^P + E_E^T\right)\right) \\
&\quad - \beta_{LH}^T S_L^T \left(I_H^P + I_H^T + \sigma\left(E_H^P + E_H^T\right)\right) + \kappa\tau S_L^J - \kappa S_L^T \\
\frac{dE_L^T}{dt} &= \beta_{LL}^T S_L^T \left(I_L^P + I_L^T + \sigma\left(E_L^P + E_L^T\right)\right) + \beta_{LE}^T S_L^T \left(I_E^P + I_E^T + \sigma\left(E_E^P + E_E^T\right)\right) \\
&\quad + \beta_{LH}^T S_L^T \left(I_H^P + I_H^T + \sigma\left(E_H^P + E_H^T\right)\right) - (\gamma + \hat{\gamma}) E_L^T + \kappa\tau E_L^J - \kappa E_L^T \\
\frac{dI_L^T}{dt} &= \gamma E_L^T - \delta I_L^T + \kappa\tau I_L^J - \kappa I_L^T \\
\frac{dR_L^T}{dt} &= \kappa\tau R_L^J - \kappa R_L^T + \delta I_L^T + \hat{\gamma} E_L^T \\
\frac{dS_E^T}{dt} &= -\beta_{EL}^T S_E^T \left(I_L^P + I_L^T + \sigma\left(E_L^P + E_L^T\right)\right) - \beta_{EE}^T S_E^T \left(I_E^P + I_E^T + \sigma\left(E_E^P + E_E^T\right)\right) \\
&\quad - \beta_{EH}^T S_E^T \left(I_H^P + I_H^T + \sigma\left(E_H^P + E_H^T\right)\right) + \kappa\tau S_E^J - \kappa S_E^T \\
\frac{dE_E^T}{dt} &= \beta_{EL}^T S_E^T \left(I_L^P + I_L^T + \sigma\left(E_L^P + E_L^T\right)\right) + \beta_{EE}^T S_E^T \left(I_E^P + I_E^T + \sigma\left(E_E^P + E_E^T\right)\right) \\
&\quad + \beta_{EH}^T S_E^T \left(I_H^P + I_H^T + \sigma\left(E_H^P + E_H^T\right)\right) - (\gamma + \hat{\gamma}) E_E^T + \kappa\tau E_E^J - \kappa E_E^T \\
\frac{dI_E^T}{dt} &= \gamma E_E^T - \delta I_E^T + \kappa\tau I_E^J - \kappa I_E^T \\
\frac{dR_E^T}{dt} &= \kappa\tau R_E^J - \kappa R_E^T + \delta I_E^T + \hat{\gamma} E_E^T \\
\frac{dS_H^T}{dt} &= -\beta_{HL}^T S_H^T \left(I_L^P + I_L^T + \sigma\left(E_L^P + E_L^T\right)\right) - \beta_{HE}^T S_H^T \left(I_E^P + I_E^T + \sigma\left(E_E^P + E_E^T\right)\right) \\
&\quad - \beta_{HH}^T S_H^T \left(I_H^P + I_H^T + \sigma\left(E_H^P + E_H^T\right)\right) + \kappa\tau S_H^J - \kappa S_H^T \\
\frac{dE_H^T}{dt} &= \beta_{HL}^T S_H^T \left(I_L^P + I_L^T + \sigma\left(E_L^P + E_L^T\right)\right) + \beta_{HE}^T S_H^T \left(I_E^P + I_E^T + \sigma\left(E_E^P + E_E^T\right)\right) \\
&\quad + \beta_{HH}^T S_H^T \left(I_H^P + I_H^T + \sigma\left(E_H^P + E_H^T\right)\right) - (\gamma + \hat{\gamma}) E_H^T + \kappa\tau E_H^J - \kappa E_H^T \\
\frac{dI_H^T}{dt} &= \gamma E_H^T - \delta I_H^T + \kappa\tau I_H^J - \kappa I_H^T \\
\frac{dR_H^T}{dt} &= \kappa\tau R_H^J - \kappa R_H^T + \delta I_H^T + \hat{\gamma} E_H^T
\end{aligned}$$

Within the Jail System:

$$\frac{dS_L^J}{dt} = -\beta_{LL}^J S_L^J \left(I_L^J + \sigma E_L^J\right) - \beta_{LE}^J S_L^J \left(I_E^J + \sigma E_E^J\right) - \beta_{LH}^J S_L^J \left(I_H^J + \sigma E_H^J\right) - \beta_{LO}^J S_L^J \left(I_O^J + \sigma E_O^J\right) + \psi_J S_L^P - \kappa\tau S_L^J - \rho S_L^J + \kappa S_L^T$$

$$\frac{dE_L^J}{dt} = \beta_{LL}^J S_L^J \left(I_L^J + \sigma E_L^J\right) + \beta_{LE}^J S_L^J \left(I_E^J + \sigma E_E^J\right) + \beta_{LH}^J S_L^J \left(I_H^J + \sigma E_H^J\right) + \beta_{LO}^J S_L^J \left(I_O^J + \sigma E_O^J\right) - (\gamma + \hat{\gamma})\, E_L^J + \psi_J E_L^P - \kappa\tau E_L^J - \rho E_L^J + \kappa E_L^T$$

$$\frac{dI_L^J}{dt} = \gamma E_L^J - \delta I_L^J - \omega_L \zeta I_L^J - (1-\zeta)\,\delta_{Death_U} I_L^J + \psi_J I_L^P - \kappa\tau I_L^J - \rho I_L^J + \kappa I_L^T$$

$$\frac{dM_L^J}{dt} = \omega_L \zeta I_L^J - \delta_{Discharge} M_L^J - \delta_{Death} \nu M_L^J$$

$$\frac{dR_L^J}{dt} = \delta I_L^J + \hat{\gamma} E_L^J + \delta_{Discharge} M_L^J + \delta_{Death} \nu M_L^J + \delta_{Death_U}(1-\nu_U)(1-\zeta) I_L^J + \delta_{Discharge}\nu_U\,(1-\zeta)\, I_L^J + \psi_J R_L^P - \kappa\tau R_L^J - \rho R_L^J + \kappa R_L^T$$

$$\frac{dS_E^J}{dt} = -\beta_{EL}^J S_E^J \left(I_L^J + \sigma E_L^J\right) - \beta_{EE}^J S_E^J \left(I_E^J + \sigma E_E^J\right) - \beta_{EH}^J S_E^J \left(I_H^J + \sigma E_H^J\right) - \beta_{EO}^J S_E^J \left(I_O^J + \sigma E_O^J\right) + \psi_J S_E^P - \kappa\tau S_E^J - \rho S_E^J + \kappa S_E^T$$

$$\frac{dE_E^J}{dt} = \beta_{EL}^J S_E^J \left(I_L^J + \sigma E_L^J\right) + \beta_{EE}^J S_E^J \left(I_E^J + \sigma E_E^J\right) + \beta_{EH}^J S_E^J \left(I_H^J + \sigma E_H^J\right) + \beta_{EO}^J S_E^J \left(I_O^J + \sigma E_O^J\right) - (\gamma + \hat{\gamma})\, E_E^J + \psi_J E_E^P - \kappa\tau E_E^J - \rho E_E^J + \kappa E_E^T$$

$$\frac{dI_E^J}{dt} = \gamma E_E^J - \delta I_E^J - \omega_H \zeta I_E^J - (1-\zeta)\,\delta_{Death_U} I_E^J + \psi_J I_E^P - \kappa\tau I_E^J - \rho I_E^J + \kappa I_E^T$$

$$\frac{dM_E^J}{dt} = \omega_H \zeta I_E^J - \delta_{Discharge} M_E^J - \delta_{Death} \nu_H M_E^J$$

$$\frac{dR_E^J}{dt} = \delta I_E^J + \hat{\gamma} E_E^J + \delta_{Discharge} M_E^J + \delta_{Death} \nu_H M_E^J + \delta_{Death_U}(1-\nu_{UH})(1-\zeta) I_E^J + \delta_{Discharge}\nu_{UH}\,(1-\zeta)\, I_E^J + \psi_J R_E^P - \kappa\tau R_E^J - \rho R_E^J + \kappa R_E^T$$

$$\frac{dS_H^J}{dt} = -\beta_{HL}^J S_H^J \left(I_L^J + \sigma E_L^J\right) - \beta_{HE}^J S_H^J \left(I_E^J + \sigma E_E^J\right) - \beta_{HH}^J S_H^J \left(I_H^J + \sigma E_H^J\right) - \beta_{HO}^J S_H^J \left(I_O^J + \sigma E_O^J\right) + \psi_J S_H^P - \kappa\tau S_H^J - \rho S_H^J + \kappa S_H^T$$

$$\frac{dE_H^J}{dt} = \beta_{HL}^J S_H^J \left(I_L^J + \sigma E_L^J\right) + \beta_{HE}^J S_H^J \left(I_E^J + \sigma E_E^J\right) + \beta_{HH}^J S_H^J \left(I_H^J + \sigma E_H^J\right) + \beta_{HO}^J S_H^J \left(I_O^J + \sigma E_O^J\right) - (\gamma + \hat{\gamma})\, E_H^J + \psi_J E_H^P - \kappa\tau E_H^J - \rho E_H^J + \kappa E_H^T$$

$$\frac{dI_H^J}{dt} = \gamma E_H^J - \delta I_H^J - \omega_H \zeta I_H^J - (1-\zeta)\,\delta_{Death_U} I_H^J + \psi_J I_H^P - \kappa\tau I_H^J - \rho I_H^J + \kappa I_H^T$$

$$\frac{dM_H^J}{dt} = \omega_H \zeta I_H^J - \delta_{Discharge} M_H^J - \delta_{Death} nu_H M_H^J$$

$$\begin{aligned}\frac{dR_H^J}{dt} &= \delta I_H^J + \hat{\gamma} E_H^J + \delta_{Discharge} M_H^J + \delta_{Death} nu_H M_H^J \\ &+ \delta_{Death_U} (1 - \nu_{UH}) (1 - \zeta) I_H^J + \nu_{UH} (1 - \zeta) \delta_{Discharge} I_H^J + \ \psi_J R_H^P \\ &- \ \kappa \tau R_H^J - \rho R_H^J + \kappa R_H^T\end{aligned}$$

$$\begin{aligned}\frac{dS_O^J}{dt} &= -\beta_{OL}^J S_O^J \left(I_L^J + I_E^J + I_H^J + \sigma \left(E_L^J + E_E^J + E_H^J\right)\right) - \beta_{OO}^J S_O^J \left(I_O^J + \sigma E_O^J\right) + \mu_J S_O^C \\ &- \mu_C S_O^J\end{aligned}$$

$$\begin{aligned}\frac{dE_O^J}{dt} &= \beta_{OL}^J S_O^J \left(I_L^J + I_E^J + I_H^J + \sigma \left(E_L^J + E_E^J + E_H^J\right)\right) + \beta_{OO}^J S_O^J \left(I_O^J + \sigma E_O^J\right) \\ &+ \mu_J E_O^C - \mu_C E_O^J - (\gamma + \hat{\gamma}) E_O^J\end{aligned}$$

$$\frac{dI_O^J}{dt} = \mu_J I_O^C - \mu_C I_O^J + \gamma E_O^J - \delta I_O^J - \omega_L I_O^J$$

$$\frac{dR_O^J}{dt} = \mu_J R_O^C - \mu_C R_O^J + \delta I_O^J + \hat{\gamma} E_O^J$$

**SI Appendix 2.** In order to determine model sensitivity to parameter values that could not be estimated directly from published/reported data, we performed a Latin hypercube sensitivity analysis (*49*). We arbitrarily selected the range of the values sampled for each parameter to be $[0.9X, 1.1X]$ where $X$ the value reported for that parameter in Tables 1 and 2. Unsurprisingly, as with most epidemic models, this model is most sensitive to parameters that govern the duration and intensity of infectivity (i.e. $\sigma$, $\gamma$ & $\hat{\gamma}$ and $\delta$). The model was also relatively insensitive to changes in the assumed increased rates of mixing among incarcerated persons in either jail or processing, suggesting that so long as there is an increase in mixing between incarcerated persons due to the structure of jails there will remain the risk of an infectious disease outbreak, rather than this being a feature of a particular combination of parameter values.

| | $I_{Com}$ | $I_{Inc}$ | $I_{Sta}$ |
|---|---|---|---|
| $\sigma$ | 0.4185 | −0.0208 | * |
| $\gamma$ | 0.2417 | 0.8015 | 0.7345 |
| $\hat{\gamma}$ | −0.3932 | −0.6453 | −0.7362 |
| $\omega$ | * | −0.0284 | * |
| $\omega_H$ | 0.0324 | −0.0370 | * |
| $\delta$ | −0.7796 | 0.0693 | * |
| $\delta_{Discharge}$ | 0.0620 | −0.0388 | * |
| $\delta_{Death}$ | 0.0594 | −0.0409 | * |
| $\delta_{DeathU}$ | 0.0613 | −0.0402 | * |
| $\nu$ | 0.0588 | −0.0430 | * |
| $\nu_H$ | 0.0588 | −0.0425 | * |
| $c_J$ | 0.0671 | 0.0702 | * |
| $c_P$ | 0.1167 | * | * |

Table 4: The sensitivity of total infections (in the Community, $I_{Com}$, in Incarcerated People, $I_{Inc}$, or in the Jail Staff, $I_{Sta}$) after 180 days in the "Shelter in Place" scenario to perturbation of each of the parameters. For completeness, these calculations include all individuals who are ever asymptomatically infected, even if they never progress to symptomatic infection. Entries marked $^*$ less than 0.01 in absolute value.

**SI Appendix 3.** The exponential survival model fit to Hu et al., 2020 estimated a mean asymptomatic shedding period of 11.79 days (95% CI: 7.86, 18.61), which is indeed a higher estimate than that found in the original manuscript, which did not account for censoring. As 5.1 of those days are already accounted for in the original estimate for $\gamma^{-}1$, this yielded an estimate for $\hat{\gamma}^{-1}$ of 6.7 days. Compared to a Kaplan-Meier fit of the available data, the exponential model fit well, and adequately models underlying survival function (Fig 10).

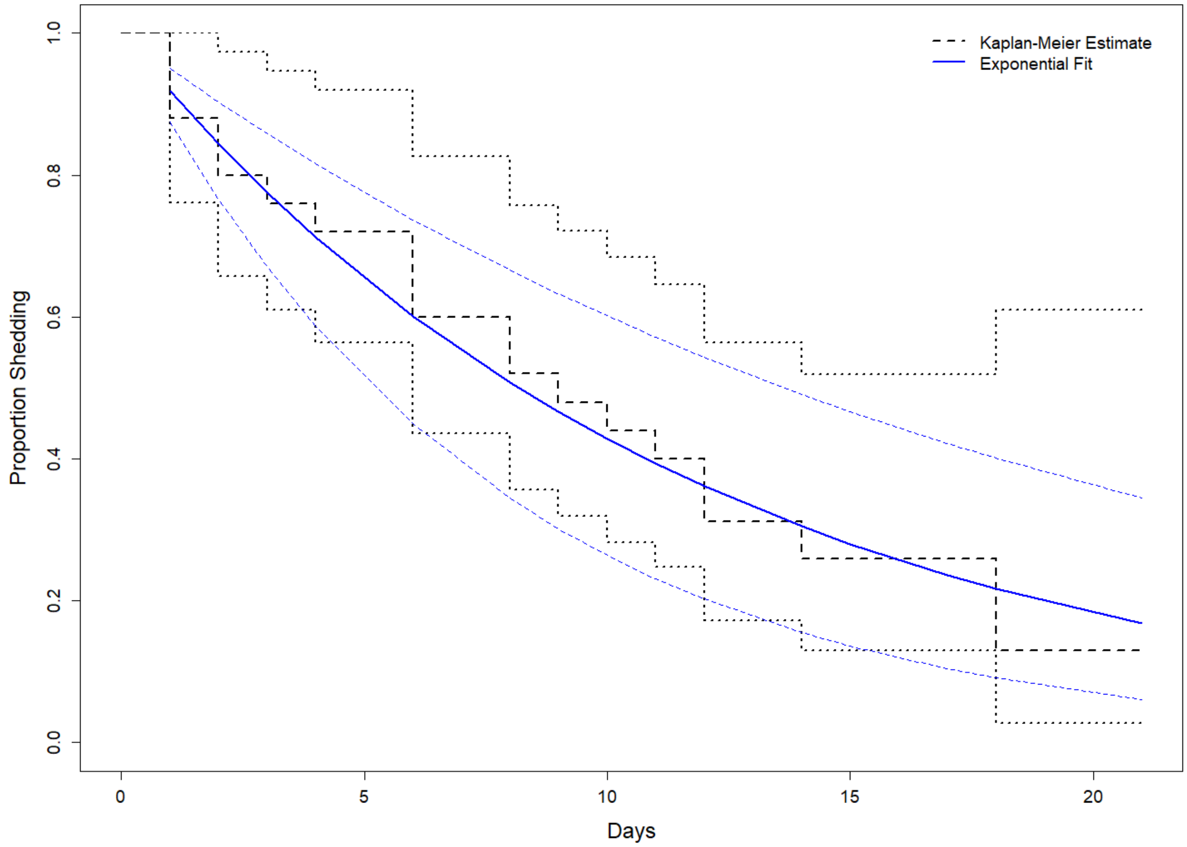


**Fig. 10.** Proportion of patients with viral shedding among a cohort of asymptomatic COVID-19 patients in China. Original data from Hu et al., 2020. Dashed black line depicts a non-parameteric Kaplan-Meier fit, while the solid blue line depicts the fit of an exponential survival function.
